# Efficacy of an Anti-Inflammatory Dietary Pattern on Global Functioning, Gut Microbiome, and Health in Patients with Psychiatric Disorders and Neurodegenerative Diseases: Protocol for a Randomized Controlled Crossover Trial

**DOI:** 10.64898/2026.06.02.26354674

**Authors:** Sophie M. van Zonneveld, Toon A.W. Scheurink, Greetje Huisman, Ellen J. van den Oever, Jasper O. Nuninga, Lisette C.P.G.M. de Groot, Teus van Laar, Benno C.M Haarman, Iris E.C. Sommer

## Abstract

**Background:** Psychiatric disorders and neurodegenerative diseases, including bipolar disorder (BD), schizophrenia spectrum disorders (SSD), Parkinson’s disease (PD), and Alzheimer’s disease (AD), are associated with substantial impairments in functioning and quality of life. Increasing evidence suggests that low-grade systemic inflammation and gut microbiome dysregulation are shared mechanisms across these brain disorders, providing a rationale for transdiagnostic interventions targeting the gut–brain axis.

**Objective:** This study was designed to evaluate the efficacy of an anti-inflammatory dietary pattern (AIDP), termed the BrAIN diet, on global functioning and a comprehensive set of secondary clinical, cognitive, inflammatory, and gut-health outcomes across relevant patient populations.

**Methods:** We designed an open-label, randomized controlled, two-period crossover trial with 12-week intervention periods, a 24-week washout period and 12-week follow-up. We aimed to enrol 100 adult outpatients (25 per diagnosis: BD, SSD, PD, and AD) aged 18–80 years, recruited through outpatient clinics and patient organisations in the northern Netherlands. Participants were randomized 1:3 to either start the BrAIN diet immediately (Group 1, BrAIN/diet-as-usual [DaU] sequence) or after 36-weeks (Group 2, DaU/BrAIN sequence). The BrAIN diet is based on Shivappa’s Dietary Inflammatory Index, components of the MIND diet, and Dutch dietary guidelines, and is delivered through weekly home-delivered food boxes, recipes, and weekly dietitian counselling. The primary outcome is global functioning measured with the Outcome Questionnaire-45 (OQ-45). The treatment effect is estimated from the timepoint × treatment interaction in a linear mixed-effects model that uses all observed timepoints, with participant as a random intercept and period and sequence as fixed effects. Secondary outcomes include Global Assessment of Functioning (GAF), cognition (Brief Assessment of Cognition, Stroop, Trail Making), quality of life (EQ-5D), fatigue (FSS), gastrointestinal symptoms (GSRS), gut-permeability biomarkers, faecal microbiome composition, inflammatory and metabolic markers, and disease-specific symptom scales. Assessments occur at weeks 0 (V1, baseline period 1), 12 (V2, end of period 1), 24 (V3, mid-washout), 36 (V4, baseline period 2), 48 (V5, end of period 2), and 60 (V6, follow-up). The trial protocol was developed in 2021 and approved by the accredited Medical Research Ethics Committee on 11 January 2022. The trial is reported in accordance with the SPIRIT 2013 guideline in effect at the time of protocol development.

**Results:** The trial received favourable ethical opinion from Medical Research Ethics Committee BeBo Assen (NL78755.056.21) on 11 January 2022 and was registered prospectively at OMON (NL-OMON52339). Recruitment commenced in February 2022; the first participant was enrolled on 7 March 2022 and the last on 6 May 2024. Follow-up was completed on 5 September 2025. A total of 107 participants were enrolled. Data analysis is ongoing; primary results are expected to be submitted for publication in summer 2026.

**Conclusions:** This study provides evidence on whether an anti-inflammatory dietary intervention targeting shared inflammatory and gut–microbiome pathways can improve global functioning and a broad set of clinical and mechanistic outcomes in psychiatric and neurodegenerative populations. The crossover design ensures all participants ultimately receive the intervention while serving as their own controls, maximising statistical power within a heterogeneous patient population. If effective, the BrAIN diet could provide a safe, accessible adjunct to standard care in neuropsychiatric and neurodegenerative populations.

**Trial Registration:** OMON NL-OMON52339; https://onderzoekmetmensen.nl/en/trial/52339

## INTRODUCTION

### Background and rationale

Psychiatric disorders and neurological diseases place a substantial burden on patients, caregivers, and society [1]. Among the most disabling are bipolar disorder (BD) and schizophrenia spectrum disorders (SSD) whitin psychiatry and Parkinson’s disease (PD) and Alzheimer’s disease (AD) among neurodegenerative diseases. Despite differing aetiologies, these four brain disorders share a recognisable clinical profile of cognitive dysfunction, fatigue, and low mood - symptoms that remain inadequately addressed by current treatments. They also share underlying biological features, most notably chronic low-grade neuroinflammation and gut-microbiome alterations, suggesting that approaches targeting these shared mechanisms could benefit patients across diagnostic boundaries.

#### NEUROINFLAMMATION AS A SHARED PATHWAY

Subtle neuroinflammation and systemic inflammation have been demonstrated in both psychiatric [2] and neurodegenerative disorders [3]. Brain-resident immune cells, particularly microglia, play essential roles in brain health through environmental sensing, neuronal support, and neuroprotective responses to pathogenic intrusion. When chronically over-activated, however, microglial activation drive prolonged neuroinflammation and increased production of pro-inflammatory cytokines including IL-1β, IL-6, TNF-α, and reactive oxygen species, with detrimental effects on brain function [3–5].

Across the four conditions, this neuroinflammatory state takes somewhat different forms. In neuropsychiatric disorders (BD and SSD), elevated immune-cell counts and increased expression of immune-related genes create a state of heightened immunoreactivity [2]; this involves not only microglia but also lymphocytes, with increased brain infiltration of B- and T-cells reported relative to healthy controls [6]. In neurodegenerative disorders (PD and AD), chronic microglial activation interacts with disease-specific protein aggregation - α-synuclein in PD and amyloid-β and tau in AD - producing self-reinforcing cycles of neuroinflammation and neuronal damage [7–8].

#### THE GUT-BRAIN AXIS IN BRAIN DISORDERS

Beyond inflammation, all four brain disorders also show altered gut-microbiome composition compared with healthy controls [9–12], pointing to a second shared mechanism. The human gut harbours a complex microbial ecosystem—the gut microbiota—comprising bacteria, archaea, viruses, fungi, and other microorganisms that together contain many times more genes than the human genome itself [13]. This microbial community has co-evolved with the host and contributes to digestion, metabolism, immune regulation, and protection against pathogens [13–14]. Bacteria are by far the most abundant and best-studied component, with the adult gut dominated by the phyla Bacteroidetes and Firmicutes [15].

Beyond its local effects in the gut, the microbiota communicates bidirectionally with the central nervous system through what is collectively termed the gut–brain axis. Three major routes of communication have been described: (i) an immune route, in which microbial signals shape systemic and central inflammatory tone via cytokines and immune cell activation; (ii) a metabolic route, in which microbial metabolites such as short-chain fatty acids (SCFAs), tryptophan derivatives, and bile acids cross the intestinal barrier and act on the brain; and (iii) a neural route, primarily via the vagus nerve and the enteric nervous system [16]. A central concept in this axis is intestinal barrier integrity: dysbiosis can compromise the gut wall (“leaky gut”), allowing microbial products such as lipopolysaccharide (LPS) - an endotoxin from Gram-negative bacteria - to enter the systemic circulation. The resulting low-grade inflammation can in turn impair the blood–brain barrier, facilitating peripheral inflammatory signals to reach the brain [17]. Disruption of microbial composition (dysbiosis) is increasingly recognised across inflammatory and neuropsychiatric conditions, providing a mechanistic basis for dietary interventions targeting brain health [18].

Among the factors that shape this microbial community, diet is one of the most important modifiable determinants of gut microbiota composition and function. High intake of animal products, processed foods, alcohol, and sugar is linked to inflammatory bacterial activity and to chronic inflammatory diseases [19]. Conversely, plant-based foods and fish are associated with beneficial bacterial species and anti-inflammatory activity [19–20]. Dietary patterns rich in nuts, fatty fish, fruits, vegetables, and whole grains promote SCFA-producing bacteria, which help control inflammation and protect epithelial integrity [19,21].

These dietary effects on the microbiota are particularly relevant for brain health, because multiple studies have demonstrated links between brain disorders and gut microbiome alterations. The kynurenine pathway – a major route of tryptophan catabolism with immunomodulatory and neuroactive effects – is disrupted in many psychiatric and neurological disorders [22–23]. Tryptophan itself, an essential amino acid and precursor of serotonin, melatonin, and vitamin B3, becomes dysregulated [24]. Recent research suggests that nutrition can modulate tryptophan metabolism, potentially decreasing neuroinflammation and improving tissue regeneration via the gut–brain axis [25].

#### DISEASE-SPECIFIC GUT-BRAIN INTERACTIONS

Within each of the four conditions, distinct patterns of gut–brain disruption have been described. In BD, systematic reviews report reduced α-diversity and depletion of butyrate-producing taxa, most consistently *Faecalibacterium* and members of the Ruminococcaceae, alongside elevations in pro-inflammatory genera, with the magnitude of dysbiosis correlating with mood-symptom severity, cognitive impairment, and fatigue [10, 26]. In SSD, dysbiosis is similarly characterised by reduced microbial diversity and disturbed SCFA production, which converge on neuroinflammation and altered microglial activity; recent meta-analyses and translational studies have positioned the gut microbiome as a candidate treatment target, including through dietary modulation [9, 27–28]. In PD, gut involvement is now considered an early — possibly initiating — feature of the disease: constipation often precedes motor symptoms by years, α-synuclein aggregation has been demonstrated in the enteric nervous system, suggesting that pathology may ascend from gut to brain via the vagus nerve (Braak’s “gut-first” hypothesis). An overabundance of pro-inflammatory and mucin-degrading bacteria (e.g., *Akkermansia,* which thins to protective intestinal mucus layer) together with depletion of SCFA-producers correlate with symptom severity [12, 29]. In AD, gut dysbiosis is associated with increased intestinal permeability and translocation of bacterial endotoxin (lipopolysaccharide, LPS) and amyloid into the circulation; resulting low-grade systemic inflammation crosses a compromised blood–brain barrier, where it amplifies microglial activation and amyloid-β deposition. [11, 30].

Although the specific microbial signatures and dominant pathways differ, the four conditions converge on a shared theme: dysbiosis-driven loss of beneficial metabolites (notably SCFAs), increased gut permeability, and a pro-inflammatory state that reaches the brain. This convergence motivates dietary interventions that target the gut microbiome as a shared therapeutic substrate across these conditions.

#### TARGETING THE GUT MICROBIOME WITH AN ANTI-INFLAMMATORY DIETARY PATTERN

Given these shared inflammatory and microbial disturbances, dietary modulation is a logical therapeutic avenue. Foods can exert pro- or anti-inflammatory effects depending on the cytokine responses they generate [19,31]. Fiber-rich products demonstrate anti-inflammatory properties [32–34], whereas processed foods, refined sugars, and alcohol promote inflammatory responses. Plant-based dietary patterns characterized by high alpha diversity and beneficial species including *Faecalibacterium prausnitzii* and *Eubacterium rectale* are associated with reduced pro-inflammatory cytokine production [19,35].

However, translating dietary evidence into benefit for these patient populations is not straightforward. People living with psychiatric disorders or neurodegenerative disease, on average, have a poorer-quality diet than age matched peers [36–37]. Common symptoms such as neuropsychological disturbances, reduced mobility, altered appetite and disrupted daily routines, interfere with their capacity to plan, prepare, and maintain a healthy dietary pattern. Dietary interventions in these populations therefore require structural support to be feasible, motivating an approach in which meals are delivered to participants’ homes and adherence is supported through weekly professional contact.

Building on this rationale, our anti-inflammatory dietary pattern (AIDP), termed the BrAIN diet [38], integrates three evidence-based components: Shivappa’s dietary inflammatory index [33, 39], components of the Mediterranean-DASH Intervention for Neurodegenerative Delay (MIND diet) shown to be beneficial for neurodegenerative diseases [40], and Dutch dietary guidelines [41–42]. This combined approach emphasizes anti-inflammatory foods while minimizing pro-inflammatory components, with the aim of optimizing gut microbiome composition and reducing systemic inflammation.

#### PRIOR WORK: DIETARY INTERVENTIONS IN BRAIN DISORDERS

Despite consistent observational evidence linking nutrition to psychiatric and neurological disorders, randomized controlled trials investigating anti-inflammatory dietary interventions remain limited across these patient populations.

For BD, literature reviews demonstrate that lifestyle interventions targeting dietary habits are effective. Reported benefits include reduced medication use, improved quality of life, and reduction in manic episodes. Despite study limitations including small sample sizes and only six available intervention studies, the beneficial effects of improving dietary habits show promise [43].

Similarly limited evidence exists for SSD. In a single dietary intervention study, Adamowicz reported that schizophrenia patients with metabolic syndrome who followed a Mediterranean-style diet - characterized by high intake of fruits, vegetables, olive oil and fish with low intake of meat and dairy products, closely resembling our BrAIN diet – showed significantly improved cognitive functioning after three months, whereas controls did not [44].

In neurodegenerative diseases, observational evidence is more substantial. For PD, Mediterranean diet adherence is associated with reduced incidence and slower progression [45] and a small intervention study reported improvements in executive function, language, attention, memory, and overall cognition [46]. For AD, Mediterranean diet adherence is similarly associated with lower incidence and slower progression [40, 45] and with lower AD biomarker scores [47]. These findings support the therapeutic potential of anti-inflammatory dietary patterns across brain disorders but underscore the need for rigorous randomized controlled trials in neuropsychiatric populations.

### Study aims and hypothesis

This exploratory trial investigates the effect of an anti-inflammatory dietary pattern on global functioning, cognitive functioning, quality of life, gut health and general health in patients with BD, SSD, PD and AD using a randomized controlled crossover design. By targeting the gut-brain axis with the BrAIN diet, we hypothesise direct effects on global functioning as brain homeostasis and plasticity improve. Reducing systemic inflammation is expected to improve mood, cognition, and quality of life across all patient groups, and the dietary pattern is also expected to reduce common gastrointestinal symptoms [20, 48].

**Primary Hypothesis:** The BrAIN diet will significantly improve global functioning (OQ-45) compared with the control condition across the four brain-disorder groups, by reducing systemic inflammation and optimising gut–brain axis communication.

**Secondary Hypotheses:** The BrAIN diet will (1) improve cognition, well-being, and fatigue; (2) reduce inflammatory markers and improve gut microbiome diversity; (3) decrease gastrointestinal symptoms and improve metabolic-health parameters; and (4) reduce disease-specific symptom severity. The intervention is expected to be safe and well-tolerated.

#### Objectives

#### PRIMARY OBJECTIVE

To investigate the effect of the BrAIN diet on global functioning measured with the Outcome Questionnaire-45 (OQ-45), with secondary assessment using the Global Assessment of Functioning (GAF) and the Individual Recovery Outcomes Counter (I.ROC), in patients with BD, SSD, PD, and AD.

#### SECONDARY OBJECTIVES

To evaluate the effect of the BrAIN diet on:

- **Psychological functioning**: Quality of life (EQ-5D) and fatigue (Fatigue Severity Scale).
- **Cognitive functioning:** Brief Assessment of Cognition, Stroop Task, and Trail Making Task.
- **Inflammation/oxidative stress:** high sensitivity-C-reactive Protein (hs-CRP), triglycerides, high density lipoprotein (HDL), free serum thiols (to reflect redox status) in blood
- **Immunological parameters:** measured by IL-6, TNF-α in blood.
- **Gut health:** gastrointestinal symptoms (Gastrointestinal Symptom Rating Scale), stool consistency (Bristol Stool Chart), intestinal permeability biomarkers (LBP, sCD14, zonulin) in blood, and faecal microbiome composition and metabolomics (faecal pH, SCFAs, redox potential).
- **General and metabolic health**: BMI, physical activity (Baecke Questionnaire), and

metabolic syndrome features (waist and hip circumference, blood pressure, glucose and triglycerides) as defined by the American Heart Association/National Heart, Lung and Blood Institute, plus medication-use monitoring.

- **Disease-specific symptoms:** Brief Psychiatric Rating Scale (BPRS); Movement Disorders Society Unified Parkinson Disease Rating Scale part III (MDS-UPDRS Part III) and Non-Motor Symptom Questionnaire (NMSQ); Instrumental Activity of Daily Living Questionnaire (IADL).
- **Stress and resilience (only in BD and SSD)\***: Childhood Trauma Questionnaire, Brugha List of Threatening Experiences, Perceived Stress Scale-10, Brief Resilience Scale, and resting state heart rate variability (HRV) measurement using ECG.
- **Oral health\***: oral health status and oral microbiome (questionnaires, intra-oral photographs, oral swab)
- **Eating behavior (PILOT, only in BD and SSD)\***: Mandometer digital scale, particularly examining alterations related to gut microbiome and antipsychotic medications.

**\*** Stress/resilience, oral health, and Mandometer assessments were added through approved protocol amendments and were not part of the original protocol (see Protocol Amendments).

## METHODS

### Study design

This is an open-label, randomized controlled, two-period crossover trial with 12-week intervention periods and a 24-week washout period (**Figure 1**). The trial framework is exploratory. Participants are randomized 1:3 to minimise carryover effects: 25% starts the BrAIN diet immediately (Group 1; BrAIN/DaU sequence), and 75% start after a 36-week delay (Group 2; DaU/BrAIN sequence). Each participant therefore undergoes a 12-week BrAIN diet condition and a 12-week control condition during which they continue their habitual diet (diet-as-usual; DaU); the two 12-week conditions are separated by the 24-week washout period and provide the within-participant contrast for the primary analysis. Outside these two assessment periods (i.e. during washout and follow-up), participants likewise follow their habitual diet, but these intervals are not part of the within-participant contrast. The trial is reported in accordance with the SPIRIT 2013 guideline, the version in effect at the time of protocol development in 2021 [49]. This protocol paper is reported following completion of recruitment and data collection, reflecting the version approved by the accredited Medical Research Ethics Committee on 11 January 2022 (NL78755.056.21). All subsequent modifications are detailed under *Protocol amendments*. The trial was prospectively registered at OMON (NL-OMON52339) prior to enrolment of the first participant; the SPIRIT checklist is provided in **Supplementary Table S1**, and the SPIRIT Schedule of Enrolment, Interventions, and Assessments figure is provided as **Table 1**.

**Figure 1.**
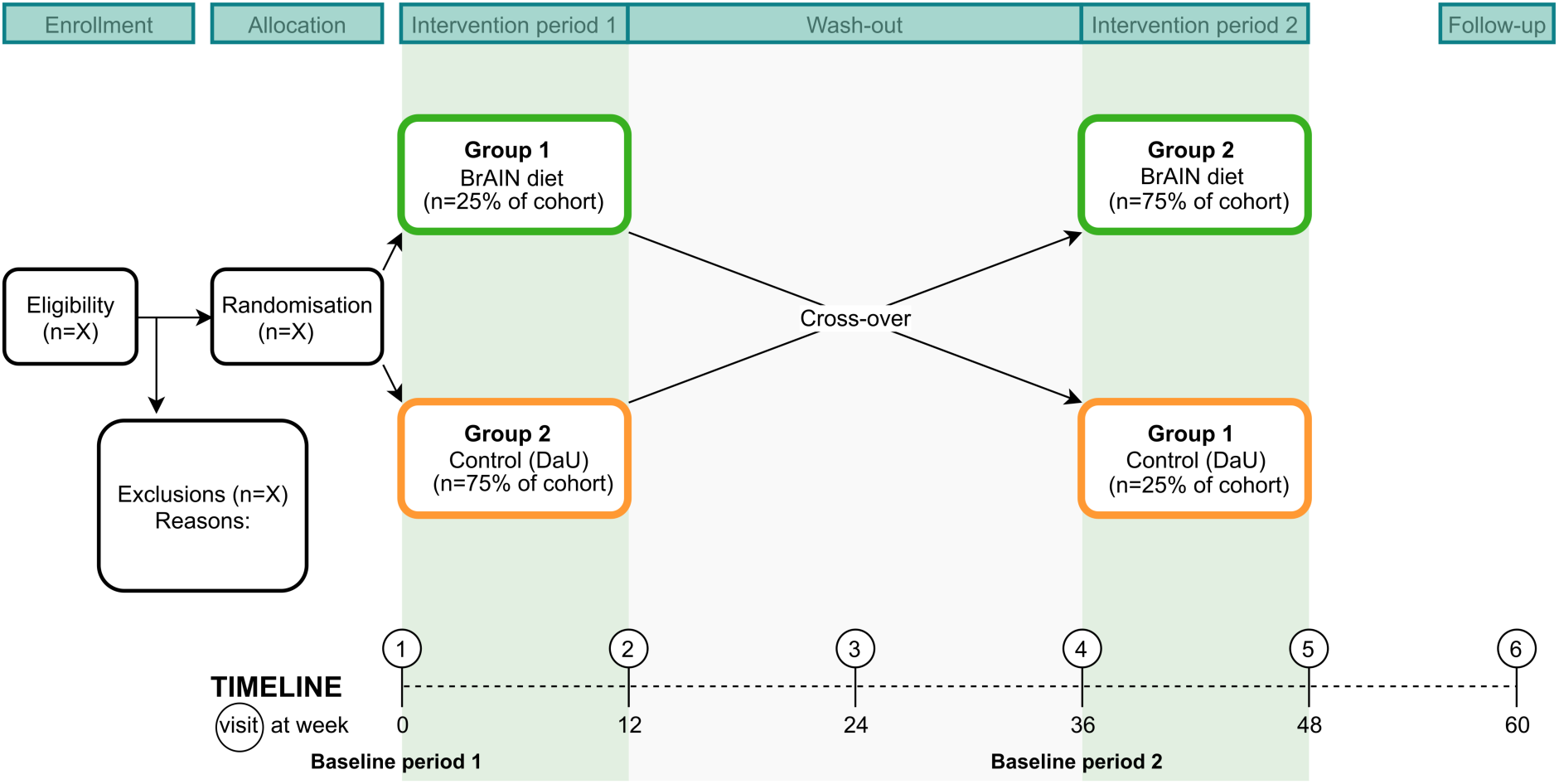
Schematic overview of the No Guts, No Glory (NGNG) trial. Eligible participants were randomised in a 1:3 ratio to two treatment sequences: Group 1 received the BrAIN diet during intervention period 1 (weeks 0–12) and the control condition during intervention period 2 (weeks 36–48), whereas Group 2 received the conditions in the reverse order. The two intervention periods were separated by a 24-week washout (weeks 12–36). Outcome assessments took place at six visits: V1 (week 0, baseline period 1), V2 (week 12, end of period 1), V3 (week 24, mid-washout), V4 (week 36, baseline period 2), V5 (week 48, end of period 2), and V6 (week 60, follow-up). The control condition was diet-as-usual (DaU), in which participants continued their habitual diet without intervention.

**Table 1.** Schedule of enrolment, interventions, and assessments for the No Guts No Glory (NGNG) trial. - an open-label, 1:3 randomised, two-period crossover trial of the BrAIN diet versus diet as usual (DaU). Trial registration: NL-OMON52339.

|  | STUDY PERIOD |  |  |  |  |  |  |  |  |
| --- | --- | --- | --- | --- | --- | --- | --- | --- | --- |
|  | Enrolment | Allocation | Post-allocation |  |  |  | Close-out | During BrAIN |  |
| Visit | Screening | V1 | V2 | V3 | V4 | V5 | V6 |  | VAS |
| Week | −6 to 0 | 0 | 12 | 24 | 36 | 48 | 60 |  | 3, 6, 9 |
| ENROLMENT |  |  |  |  |  |  |  |  |  |
| Eligibility screen | X |  |  |  |  |  |  |  |  |
| Informed consent |  | X |  |  |  |  |  |  |  |
| In-/exclusion criteria |  | X |  |  |  |  |  |  |  |
| Randomisation & allocation |  | X |  |  |  |  |  |  |  |
| Re-assessment of decisional capacity |  |  |  | X |  |  |  |  |  |
| INTERVENTIONS |  |  |  |  |  |  |  |  |  |
| BrAIN/DaU sequence (≈25%) |  | X | X | X | X | X |  |  |  |
| DaU/BrAIN sequence (≈75%) |  | X | X | X | X | X |  |  |  |
| Weekly contact / dietetic support during BrAIN |  |  |  |  |  |  |  | X |  |
| Adherence monitoring (≥80% of products consumed) |  |  |  |  |  |  |  | X |  |
| BASELINE VARIABLES |  |  |  |  |  |  |  |  |  |
| Demographics, medical history, medication use | X | X |  |  |  |  |  |  |  |
| OUTCOME ASSESSMENTS |  |  |  |  |  |  |  |  |  |
| Primary outcome |  |  |  |  |  |  |  |  |  |
| Global functioning (OQ-45) |  | X | X | X | X | X | X |  |  |
| Secondary outcomes — clinical |  |  |  |  |  |  |  |  |  |
| Global functioning (GAF, I.ROC) |  | X | X | X | X | X | X |  |  |
| Quality of life (EQ-5D-5L-VAS) |  | X | X | X | X | X | X |  |  |
| Cognitive functioning (BAC, Stroop, TMT) |  | X | X |  | X | X |  |  |  |
| BD/SSD symptom severity (BPRS) |  | X | X | X | X | X | X |  |  |
| PD symptom severity (MDS-UPDRS Part III; NMSQ) |  | X | X | X | X | X | X |  |  |
| AD symptom severity (IADL) |  | X | X | X | X | X | X |  |  |
| Fatigue (FSS) |  | X | X | X | X | X | X |  |  |
| Gastrointestinal symptoms (GSRS) |  | X | X | X | X | X | X |  |  |
| Physical activity (Baecke) |  | X | X | X | X | X | X |  |  |
|  | Enrolment | Allocation | Post-allocation |  |  |  | Close-out | During BrAIN |  |
| Visit | Screening | V1 | V2 | V3 | V4 | V5 | V6 |  | VAS |
| Week | -6 to 0 | 0 | 12 | 24 | 36 | 48 | 60 |  | 3, 6, 9 |
| Stress & resilience — BD/SSD only (CTQ-SF, Brugha LTE, PSS-10, BRS) |  | X | X |  | X | X |  |  |  |
| Oral health & oral self-care (optional) |  | X | X |  | X | X |  |  |  |
| Eating behaviour — BD/SSD only (Mandometer, optional, once) |  |  | X | X | X | X | X |  |  |
| **Secondary outcomes — clinical & biospecimen** |  |  |  |  |  |  |  |  |  |
| Anthropometry (BMI, waist & hip), blood pressure |  | X | X | X | X | X | X |  |  |
| Body weight monitoring (mid-intervention, week 6) |  |  |  |  |  |  |  | X |  |
| Metabolic syndrome status (NCEP ATP III); fatty liver index |  | X | X |  | X | X |  |  |  |
| Venous blood sample (metabolic, hs-CRP, inflammation, gut-permeability biomarkers) |  | X | X |  | X | X |  |  |  |
| Faecal sample (microbiome, metabolites, calprotectin) |  | X | X |  | X | X |  |  |  |
| Stool consistency (Bristol Stool Chart) |  | X | X |  | X | X |  |  |  |
| Heart rate variability (ECG) — BD/SSD only |  | X | X |  | X | X |  |  |  |
| **DIETARY INTAKE & ADHERENCE** |  |  |  |  |  |  |  |  |  |
| Diet adherence (MIND-NL-Eetscore-FFQ) |  | X | X | X | X | X | X |  |  |
| Food Frequency Questionnaire (FFQ, recall last 4 weeks) |  |  | X |  |  | X |  |  |  |
| 5-day food diary (week 3 of each period) |  |  |  |  |  |  |  | X (/control) |  |
| 24-h dietary recall (mid-BrAIN only) |  |  |  |  |  |  |  | X |  |
| Participant experience / satisfaction (VAS 1–10) |  |  |  |  |  |  |  |  | X |
| **SAFETY MONITORING** |  |  |  |  |  |  |  |  |  |
| Adverse events — actively monitored during BrAIN (visits + weekly contacts) |  | X | X | X | X | X | X | X |  |
| GI tolerability of BrAIN diet (VAS 1–10) |  |  |  |  |  |  |  |  | X |
| Serious adverse events (reported to sponsor within 24 h) |  | X | X | X | X | X | X | X | X |
| Green cells = active BrAIN diet exposure during a given period; orange cells = diet as usual (DaU). V1 and V4 are baseline visits for intervention periods 1 and 2, respectively; V2 and V5 are post-intervention visits; V3 is a mid-study wash-out visit; V6 is the 12-week post-period-2 follow-up visit. Each intervention period lasts 12 weeks; the wash-out period between V2 and V4 lasts 24 weeks. Stratified randomisation (1:3) is performed by an independent researcher using computer-generated permuted block randomisation (Sealed Envelope), with stratification by sex and by diagnostic group. The BrAIN/DaU sequence allocates BrAIN diet first (≈25%); the DaU/BrAIN sequence allocates BrAIN diet second (≈75%). Adherence is defined as consumption of ≥80% of delivered food products during the BrAIN period, evaluated by dietary coaches using 24-h recall and ongoing contact data. The "Weekly" column denotes contact moments and assessments occurring throughout each 12-week BrAIN period; the "Wk 3, 6, 9" column denotes specific in-intervention timepoints for visual-analogue-scale (VAS) ratings. *Abbreviations:* AD, Alzheimer's disease; AE, adverse event; BAC, Brief Assessment of Cognition; BAECKE, Baecke physical activity questionnaire; BD, bipolar disorder; BPRS, Brief Psychiatric Rating Scale; BrAIN, Brain Anti-Inflammatory Nutrition diet; BRS, Brief Resilience Scale; CTQ-SF, Childhood Trauma Questionnaire–Short Form; DaU, diet as usual; ECG, electrocardiography; EQ-5D, EuroQol-5 Dimensions; EQ-5D-VAS, EuroQol-5D Visual Analogue Scale; FFQ, Food Frequency Questionnaire; FSS, Fatigue Severity Scale; GAF, Global Assessment of |  |  |  |  |  |  |  |  |  |
|  | Enrolment | Allocation | Post-allocation |  |  |  | Close-out | During BrAIN |  |
| Visit | Screening | V1 | V2 | V3 | V4 | V5 | V6 |  | VAS |
| Week | -6 to 0 | 0 | 12 | 24 | 36 | 48 | 60 |  | 3, 6, 9 |
| Functioning; GI, gastrointestinal; GSRS, Gastrointestinal Symptom Rating Scale; hs-CRP, high-sensitivity C-reactive protein; HRV, heart rate variability; IADL, Instrumental Activities of Daily Living questionnaire; I.ROC, Individual Recovery Outcomes Counter; LTE, List of Threatening Experiences; MDS-UPDRS, Movement Disorder Society–Unified Parkinson's Disease Rating Scale; MIND-NL-Eetscore-FFQ, Mediterranean–DASH Intervention for Neurodegenerative Delay (Dutch adaptation) screener; NCEP ATP III, National Cholesterol Education Program Adult Treatment Panel III; NMSQ, Non-Motor Symptoms Questionnaire; OQ-45, Outcome Questionnaire-45; PD, Parkinson's disease; PSS-10, 10-item Perceived Stress Scale; SAE, serious adverse event; SSD, schizophrenia spectrum disorder; TMT, Trail Making Test; VAS, visual analogue scale. |  |  |  |  |  |  |  |  |  |

### Trial Setting

The trial is conducted at outpatient clinics in the northern Netherlands, including the University Medical Center Groningen (UMCG), Martini Hospital, care centre Maartenshof, the Parkinson Expertise Centers of Groningen and Leeuwarden, Medical Centre Leeuwarden, GGZ Lentis, and GGZ Friesland. All study visits are conducted at participants’ homes or at the UMCG. The sponsor and coordinating site is the UMCG, Groningen, The Netherlands.

### Eligibility criteria

#### INCLUSION CRITERIA

To be eligible, participate must meet all of the following criteria:

1. Clinical diagnosis made by medical specialist of bipolar disorder type 1 or type 2, schizophrenia, schizophreniform disorder, schizoaffective disorder, Parkinson’s disease or Alzheimer’s disease.
2. Living in the Dutch provinces Drenthe, Friesland or Groningen.
3. Cognitive capacity* to understand what participation entails (confirmed by clinical judgement of the medical specialist), and willing to provide written informed consent.
4. Aged 18-80 years and sufficient command of the Dutch language.
5. Motivated and capable to follow the dietary pattern (use food from deliverd boxes) and to participate in interview visits at home; partner or other household members support participation, or at least do not oppose it.
6. Able to consume foods as prescribed, without religious, medical or sociocultural factors precluding participation or adherence to the diet.
7. Living independently (not in nursing home) and is willing and able to prepare fresh meals using standard kitchen equipment.

* Legal capacity is reassessed at mid-washout period (week 24, V3). Participants who become incapacitated during the study are withdrawn.

#### EXCLUSION CRITERIA

Participants are excluded if any of the following apply:

1. Pregnancy or breastfeeding (or anticipated pregnancy during the study period).
2. Severe under- or overweight requiring medical treatment (assessed by a gastrointestinal specialist).
3. Severe bowel or liver disease, or acute/chronic pancreatitis (assessed by a gastrointestinal specialist).
4. Inability to consume the delivered products exclusively (e.g., nut allergy), need for an incompatible diet (e.g., diabetes-specific diet, food intolerances), or specific dietary preferences incompatible with the intervention (e.g., vegan, vegetarian, no fish).
5. Already consuming an AIDP on own initiative (assessed with the MIND-NL-Eetscore-FFQ, threshold ≥10).
6. Current use of antibiotics (or use within the past 4 weeks); regular use of probiotics (e.g., Yakult, Vifit, Activia) or specific prebiotic supplements without willingness to discontinue at least 6 weeks before and during the trial.

### Recruitment

Participants were identified through outpatient clinics at the recruiting centres listed above. Treating physicians asked eligible patients whether researchers from the UMCG could approach them, and potential participants were also informed about the study via patient organisations and Hersenonderzoek.nl, the Dutch national patient registry for brain research. To facilitate participation, all study visits were conducted at participants’ homes by default, with the UMCG offered as alternative location and travel costs reimbursed when applicable. Multiple complementary outreach activities were undertaken throughout the recruitment period, including presentations at patient association meetings, newsletters distributed via collaborating patient organisations, repeat invitations through treating physicians, and targeted outreach via diagnosis-specific patient networks.

### Patient and public involvement

A study-specific patient advisory board, comprising individuals living with one of the studied conditions (Parkinson’s disease, bipolar disorder, schizophrenia spectrum disorder, or Alzheimer’s disease), was established to inform the design and conduct of the trial. The board provided input on the feasibility of the intervention (home visits, weekly food boxes, and dietary approach), on the participant-facing study materials (information letter, informed consent form, and brochures), and on the selection and wording of some of the questionnaires and instruments. The board met quarterly throughout the trial, allowing iterative adjustments where needed.

### Sample size

Sample size was calculated for the primary outcome (OQ-45). Assuming an expected effect size (Cohen’s d) of 0.5, α=0.05 (two-sided), and β=0.20 (80% power), 64 participants are required for the primary analysis. To allow for an anticipated dropout rate of 30%—common in dietary intervention trials—we planned to enrol 100 participants (originally 25 per diagnostic group: BD, SSD, PD, and AD). The crossover design provides up to 400 observations across time points. The primary analysis was planned at transdiagnostic level; secondary exploratory analyses were planned to compare (i) psychiatric versus neurodegenerative disorders, and (ii) individual diagnostic groups.

#### Randomisation, allocation concealment, and blinding

#### SEQUENCE GENERATION

The randomisation sequence was generated in advance using sealed envelope (sealed envelope ltd, london, uk; https://www.sealedenvelope.com/), with a 1:3 allocation ratio (Group 1: BrAIN/DaU sequence; Group 2: DaU/BrAIN sequence), permuted block randomisation (block size 4), and stratification by diagnosis and sex. Sealed envelope produced separate randomisation lists per diagnostic stratum, exported as csv files for secure storage and use by the independent investigator.

#### ALLOCATION CONCEALMENT

Allocation concealment was achieved by separating the randomisation sequence from the recruitment and assessment team. The pre-generated randomisation lists were held by an independent investigator who had no role in participant enrolment, intervention delivery, or outcome assessment, and were stored in a secure environment inaccessible to the study team. When a new participant was eligible for inclusion, the recruiting researcher provided only the participant’s diagnosis and sex; the independent investigator returned the next allocation from the corresponding stratified list, together with the participant’s sequential study identifier (NG001, NG002, …). The full sequence was not shared with the study team during recruitment; randomisation lists were only transferred to the study team after recruitment had been completed, for analysis and archival purposes.

#### IMPLEMENTATION

Sequence generation, allocation, and assignment of participants to interventions were performed by the independent investigator described above; the recruiting researchers were responsible for screening, consenting, and enrolling participants but had no access to the randomisation lists.

#### BLINDING

Because the intervention is a home-delivered dietary pattern with weekly counselling, blinding of participants and intervention providers is not feasible (open-label design). Outcome assessors are not formally blinded. However, the primary outcome (OQ-45) is participant-reported, which limits the influence of assessor knowledge on the primary endpoint. Biomarker analyses (microbiome, blood inflammation/permeability markers) are performed by laboratory personnel who do not have access to allocation information. Samples were labelled only with the participant’s coded study identifier (NG001, NG002, …) without any indication of group assignment, and the allocation key was not shared with the laboratory. Statisticians performing the analyses were not blinded to allocation.

### Intervention

#### THE BRAIN ANTI-INFLAMMATORY NUTRITIONAL (BRAIN) DIET

The BrAIN diet integrates three evidence-based components: (1) Shivappa’s Dietary Inflammatory Index [33, 39]; (2) components of the MIND diet [40]; and (3) the Dutch Healthy Diet guidelines [41–21]. The pattern emphasises high intake of fruits, vegetables, legumes, whole grains, nuts, fermented dairy, fish, and olive oil, while limiting processed foods, red and processed meat, refined sugar, and alcohol (see also **Supplementary Table S2**). Individual dietary plans are created based on participants’ energy needs through dietitian consultation. A more detailed description of the BrAIN diet has been published [38]. An example basis week-and daily menu is provided in **Supplementary Table S3**, and all dinner recipes used during the intervention are publicly available [50].

#### INTERVENTION DELIVERY

During the 12-week BrAIN intervention period, participants receive:

- Weekly home-delivered food boxes containing all necessary anti-inflammatory products.
- Weekly dietitian counselling, delivered through home visits or (video) calls, to monitor adherence and provide support.
- Recipes and instructional cooking videos.
- Practical preparation assistance where needed.

#### INTERVENTION MODIFICATIONS AND DISCONTINUATION CRITERIA

The intervention may be modified or discontinued for an individual participant in three situations: in response to an adverse event, at the participant’s request, or because of a change in clinical status. To detect such situations early, adverse events are actively monitored at every study visit and at the weekly dietitian contacts during the BrAIN intervention period; during the diet-as-usual (control) period, adverse events are recorded only at the scheduled study visits or upon spontaneous report. In addition, participants rate gastrointestinal tolerability on a visual analogue scale (VAS, 1–10) at weeks 3, 6, and 9 of the intervention. When intolerability arises, the default response is adaptation rather than discontinuation: the dietitian offers exchange of the offending food items for alternatives within the BrAIN food choice list during the next weekly consultation, so that overall adherence to the dietary pattern is preserved. Body weight is formally assessed in week 6 of the BrAIN intervention period; however, the dietitian also remains alert to early signs of clinically relevant unintended weight loss during the weekly contacts and acts before the midway assessment if such signs emerge. In either case, the dietitian reviews the participant and, where indicated, adjusts the energy density of the individualised meal plan. Participants may withdraw from the intervention at any time and for any reason, without consequences for their regular care. Beyond such voluntary withdrawal, premature discontinuation is considered when one of the following occurs: (i) a serious adverse event judged at least possibly related to the intervention, (ii) a new medical condition incompatible with continued participation, or (iii) withdrawal of informed consent. Any serious adverse event is reported to the sponsor within 24 hours.

#### ADHERENCE MONITORING

Adherence to the BrAIN dietary pattern is supported by three complementary strategies. First, all food products required for the diet are delivered weekly to participants’ homes at no cost, removing the financial and logistical barriers that often limit dietary adherence. Second, every delivery is accompanied by detailed recipes and instructional cooking videos, and the study dietitian develops an individualised meal plan based on each participant’s energy needs prior to the start of the intervention. Third, the dietitian holds weekly consultations with each participant by home visit or video call; during these consultations, participants can request exchange of specific food items within the BrAIN food choice list, discuss preparation techniques, and, where needed, receive practical preparation assistance.

Adherence is monitored using a combination of qualitative and quantitative procedures. The weekly dietitian contacts and the inspection of returned food boxes provide ongoing qualitative information on how closely each participant follows the dietary pattern. For quantitative assessment, we use the MIND-NL-Eetscore-FFQ, a brief food-frequency questionnaire adapted from the American MIND scoring system to the Dutch context; total scores range from 0 to 15, with higher scores reflecting greater adherence to the anti-inflammatory dietary pattern. At baseline, participants are required not to already follow an anti-inflammatory dietary pattern, operationalised as a MIND-NL-Eetscore <10 at screening (see Eligibility criteria). At the end of each intervention period, participants are considered adequately adherent if their MIND-NL-Eetscore is ≥10. Participants who do not meet this threshold are flagged as protocol deviations and are excluded from the per-protocol sensitivity analysis; all randomised participants are, however, retained in the primary intention-to-treat analysis. To complement the MIND-NL-Eetscore, dietary intake is characterised through three additional methods. First, a 5-day food diary is completed in week 3 of both the BrAIN diet and the control periods; participants record all foods and beverages consumed over five consecutive days, including one weekend day, using weighed records or household measures. Mean daily intake is then entered into NutriCount (version 2024) to obtain energy and nutrient values. Second, a comprehensive Food Frequency Questionnaire is administered at the post-intervention visit of both periods to characterise habitual dietary intake over the preceding four weeks. Third, during the BrAIN intervention period only, a single 24-hour dietary recall is conducted by a dietary coach at a random mid-intervention timepoint; this recall serves both to monitor compliance and to identify potential nutrient deficiencies that can be addressed through dietary adjustments at the next consultation. On the basis of the 24-hour recall data and the ongoing weekly contacts, the dietary coach rates whether each participant consumed at least 80% of the delivered food products; this constitutes an additional pre-specified compliance criterion. Finally, the plausibility of reported energy intake from the food diaries is assessed using the Goldberg method [51].

#### CONCOMITANT CARE

Standard medical care continues unchanged for all participants throughout the trial. In particular, disease-specific pharmacological treatment and regular contact with treating physicians are maintained as usual. To minimise confounding of the dietary effect, participants are asked to keep all chronic medication stable during the intervention and washout periods; medically indicated changes are nonetheless permitted and are recorded at every study visit. Because antibiotics and probiotics may directly affect the gut microbiome - a key secondary outcome - antibiotic use and regular probiotic supplementation are exclusion criteria at baseline (see Eligibility criteria). During the trial, any incidental antibiotic course or over-the-counter supplement use is recorded so that it can be considered in sensitivity analyses. Participants are also asked not to start a new structured dietary pattern, weight-loss programme, or supplementation regimen during the trial without informing the study team. Non-pharmacological care, including physiotherapy, psychotherapy, occupational therapy, and other allied-health interventions, is permitted and continues according to standard care.

#### Outcome measures

#### PRIMARY OUTCOME MEASURE

Global functioning measured with the Outcome Questionnaire 45 (OQ-45) [52], a 45-item self-report questionnaire covering symptomatic distress, interpersonal relationships, and social role problems. The analysis metric is the within-participant difference between intervention and control differential change from pre to post between the BrAIN diet and the control condition (DaU), estimated from the timepoint × treatment interaction in a linear mixed-effects model; assessment time points are weeks 0 (V1, baseline period 1), 12 (V2, end of period 1), 24 (V3, mid-washout), 36 (V4, baseline period 2), 48 (V5, end of period 2), and 60 (V6, follow-up).

#### SECONDARY OUTCOME MEASURES

- **Global functioning (additional):** Clinician-reported Global Assessment of Functioning (GAF) [53] Individual Recovery Outcomes Counter (I.ROC) [54]
- **Cognitive functioning:** Brief Assessment of Cognition (BAC) [55], Stroop Task [56], Trail Making Task [57]
- **Quality of life**: EuroQoL 5D (EQ-5D) [58–59]
- **Fatigue:** Fatigue Severity Scale (FSS) [60]
- **Gastrointestinal symptoms:** Gastrointestinal Symptom Rating Scale (GSRS) [61], Bristol Stool Chart [62]
- **General and metabolic health:** BMI, metabolic syndrome features (waist/hip circumference, blood pressure, fasting glucose, triglycerides); physical activity (Baecke questionnaire) [63]
- **Gut permeability and inflammation:** Zonulin, LPS binding protein (LBP), soluble CD14 (sCD14), I-FABP, EndocAb in blood; fecal calprotectin
- **Microbiome and metabolomics**: Shotgun metagenomic sequencing, faecal pH, SCFAs, redox potential
- **Inflammation and oxidative stress**: hs-CRP, triglycerides, HDL, free serum thiols, IL-6, TNF-α
- Disease-specific questionnaires:
- `o` Brief Psychiatric Rating Scale (BPRS) [64]
- `o` MDS-UPDRS part III [65], Non-Motor Symptom Questionnaire (NMSQ) [66]
- `o` Instrumental Activities of Daily Living (IADL) [67]
- **Acceptability and participant experience:** visual analogue scales (VAS, 1–10) at weeks 3, 6, and 9 of the BrAIN intervention period, rating satisfaction with the dinner recipes and overall satisfaction with the dietary programme

#### ADDITIONAL OUTCOME MEASURES (ADDED THROUGH AMENDEMENTS)

- **Stress and resilience (BD and SSD only):** Brief Resilience Scale [68]; Childhood Trauma Questionnaire–Short Form [69]; Perceived Stress Scale-10 [70]; Brugha List of Threatening Experiences [71]; resting-state heart rate variability (HRV) measurement (ECG).
- **Oral health (optional):** self-care questionnaire; Oral Health Impact Profile (OHIP); intra-oral photograph; oral microbiome swab.
- **Eating behaviour (BD and SSD only, optional):** Mandometer assessment of pattern and speed of food intake during a standardised meal.

#### SCHEDULE OF ASSESSMENTS

Visits take place at participants’ homes or at the UMCG. Blood and faecal samples are collected at Weeks 0 (V1), 12 (V2), 36 (V4), and 48 (V5). Cognitive testing is conducted at pre- and post-intervention visits only. The full schedule of enrolment, interventions, and assessments is provided as **Table 1.** A complete overview of assessments per visit and their duration is provided in **Supplementary S2**.

### Data collection methods

Self-report questionnaires are administered on paper or tablet at home visits, depending on participant preference and capability. Cognitive tasks (BAC, Stroop, Trail Making) are administered by trained researchers at home or at the UMCG. Blood samples are drawn by trained personnel at the UMCG or by mobile phlebotomy at participants’ homes. Faecal samples are self-collected by participants using a standardised home collection kit and returned to the laboratory. All assessors and laboratory personnel receive standardised training on study procedures, in line with Good Clinical Practice (GCP).

All trial instruments are validated questionnaires or established cognitive tests used in their previously validated versions; for Dutch-language questionnaires, the validated Dutch translation is used where available. Reliability and validity of each instrument have been demonstrated in prior validation studies, to which we refer rather than reproducing the underlying psychometric statistics. **Table 2** summarises the assessment domain, instrument, language version used in the trial, and the primary validation reference for each questionnaire and cognitive test. For instruments administered only in specific diagnostic groups, the relevant group is indicated.

**Table 2.** Trial instruments with assessment domain, language version, and validation references

| Domain | Instrument | Version | Validation reference |
| --- | --- | --- | --- |
| Global functioning (primary) | Outcome Questionnaire 45 (OQ-45) | Dutch | De Jong & Spinhoven, 2008 [52] |
| Global functioning | Global Assessment of Functioning (GAF) | Dutch; clinician-rated | Jones et al., 1995 [53] |
| Global functioning | Individual Recovery Outcomes Counter (I.ROC) | Dutch; self-report | Beckers et al., 2022 (Dutch validation) [54] |
| Cognitive functioning | Brief Assessment of Cognition (BAC) | Dutch | Keefe et al., 2004 [55] |
| Cognitive functioning | Stroop Task | Standard administration | Stroop, 1992 [56] |
| Cognitive functioning | Trail Making Test (Parts A and B) | Standard administration | Corrigan & Hinkeldey, 1987 [103] |
| Health-related quality of life | EuroQol EQ-5D-5L | Dutch | Herdman et al., 2011 [59]; EuroQol Group, 1990 [58] |
| Fatigue | Fatigue Severity Scale (FSS) | Dutch; self-report | Herlofson & Larsen, 2002 [60] |
| Gastrointestinal symptoms | Gastrointestinal Symptom Rating Scale (GSRS) | Dutch | Revicki et al., 1997 [61] |
| Stool consistency | Bristol Stool Form Scale | Standard administration | Lewis & Heaton, 1997 [62] |
| Symptom severity (BD, SSD) | Brief Psychiatric Rating Scale (BPRS) | Clinician-rated | Reference [64] |
| Functional disability (AD) | Instrumental Activities of Daily Living (IADL) | Dutch | Reference [67] |
| Motor symptoms (PD) | MDS-UPDRS Part III | Dutch; clinician-rated | Reference [65] |
| Non-motor symptoms (PD) | Non-Motor Symptoms Questionnaire (NMSQ) | Dutch; self-report | Reference [66] |
| Childhood adversity (BD, SSD) | Childhood Trauma Questionnaire – Short Form (CTQ-SF) | Dutch | Reference [69] |
| Recent stressful events (BD, SSD) | Brugha's List of Threatening Experiences | Dutch | Brugha et al., 1985 [71] |
| Perceived stress (BD, SSD) | Perceived Stress Scale, 10-item (PSS-10) | Dutch | Cohen et al., 1983 [70] |
| Resilience (BD, SSD) | Brief Resilience Scale (BRS) | Dutch | Smith et al., 2008 [68] |
| Physical activity | Baecke Questionnaire | Dutch; self-report | Reference [63] |
| Dietary adherence | MIND-NL-Eetscore-FFQ | Dutch | See Methods (Adherence Monitoring) |
| Dietary intake (habitual) | Food Frequency Questionnaire (FFQ) | Dutch (Siebelink) | Siebelink et al., 2011 |

### Retention

Strategies to promote retention include weekly dietitian contact during BrAIN intervention periods, flexibility in scheduling home visits, and reimbursement of travel costs where applicable. For participants who discontinue or deviate from the intervention, all subsequent scheduled outcome assessments are still attempted in line with the intention-to-treat principle, where consent permits.

### Data management

All study data are collected and managed in an eCRF using REDCap (Research Electronic Data Capture, version 16.0.17), a secure web-based application hosted by UMCG IT Services. The procedures described in this section constitute the data management plan for this trial. To control access and protect data integrity, each study team member receives a personal username and password, and access is granted on a role-based basis, so that investigators, dietitians, and trial monitors have different levels of access depending on their study role. All data are encrypted in transit, and REDCap automatically maintains an audit trail that records every change to a data field together with the timestamp and user identifier. To protect participant confidentiality, each participant is identified by a coded study identifier (NG followed by a sequential number, e.g., NG001). The key linking these codes to identifiable participant data is held by the research team and stored separately from the study data, in accordance with the EU General Data Protection Regulation (GDPR) and the Dutch Implementing Act (UAVG). Data quality is safeguarded through several complementary procedures. First, REDCap enforces field-level validation of data formats (e.g., dates, integers) and applies predefined range checks to flag biologically implausible values at the moment of entry; branching logic further ensures that only relevant questions are presented, and required fields prevent submission of incomplete forms. Second, although double data entry is not used, all data entered into REDCap are subsequently verified by trained research interns against the original source documents to detect entry errors. Third, REDCap’s Data Quality module is run periodically to identify missing values, outliers, and internal inconsistencies. Finally, source data verification of a random subset of records is performed by an independent trial monitor (see §Trial monitoring). All study personnel involved in data collection or entry receive standardised training in line with Good Clinical Practice. All study documents and records are retained for 15 years after the end of the study; biological samples (blood and faeces) are retained in storage for 5 years and may be used for future research only with explicit additional consent.

### Statistical analysis

All analyses are conducted in R (version 4.3.3 or later), and both intention-to-treat and per-protocol analyses are pre-specified.

#### PRIMARY ANALYSES

The effect of the BrAIN diet on the OQ-45 is estimated using a linear mixed-effects model. The outcome is modelled as a function of timepoint (pre versus post within each intervention period), treatment condition (BrAIN versus control), and their interaction (timepoint × treatment). Period and treatment sequence are included as fixed effects, and participant is included as a random intercept to account for the within-participant correlation that arises from the two-period crossover design. Age and sex are included as fixed-effect covariates. The treatment effect is estimated from the timepoint-by-treatment interaction term, which represents the differential change from pre to post between the BrAIN diet and the control condition.

We chose this modelling approach because it combines three properties that are particularly well suited to a crossover trial with repeated measurements. It yields a directly interpretable estimate of the differential change between conditions; it accommodates the within-participant correlation intrinsic to the design; and, importantly, it uses all observed timepoints rather than requiring complete pre–post pairs, so that participants with partial data still contribute information (see also Missing data, below).

Results are reported with point estimates, 95% confidence intervals, and exact P values. Model-based standardised effect sizes (Cohen’s d) are derived by dividing the estimated interaction effect by the residual standard deviation of the model.

#### CARRYOVER AND PERIOD EFFECTS

Carryover and period effects are formerly tested. If a meaningful carryover effect is detected, the primary analysis is restricted to data from the first treatment period only (parallel-group analysis); the corresponding analysis using the full crossover dataset is then also reported as a sensitivity analysis, so that both perspectives are transparently available to the reader.

#### SECONDARY ANALYSES

Secondary outcomes are analysed using the same linear mixed-effects model structure as the primary analysis, with one model per outcome. To control the false discovery rate across the family of secondary outcomes, P values are adjusted using the Benjamini–Hochberg procedure, and effect sizes are reported as Cohen’s d.

Two types of subgroup analyses are pre-specified: (i) psychiatric (BD, SSD) versus neurodegenerative (PD, AD) disorders, and (ii) per-diagnosis analyses. Because group sizes are limited, per-diagnosis analyses are interpreted as exploratory. Disease-specific symptom severity instruments (BPRS, MDS-UPDRS, NMSQ, IADL) are analysed within their respective diagnostic groups only.

#### MICROBIOME ANALYSES

Microbiome data are analysed in three complementary ways: alpha- and beta-diversity metrics across conditions; the relative abundance of pre-specified taxa (e.g., *Ruminococcaceae*); and hypothesis-driven analyses of specific genera selected on the basis of prior literature (e.g., *Prevotella*). For each, both intention-to-treat and per-protocol analyses are reported where feasible, depending on adherence and dropout. Additionally, shotgun metagenomic sequencing enables species- and strain-level resolution and functional gene profiling, which inform exploratory analyses of microbial metabolic pathways.

#### MISSING DATA

The linear mixed-effects model used for the primary and secondary analyses is robust to missing observations under the missing-at-random (MAR) assumption: participants with one or more missing measurements still contribute information through their observed timepoints, and maximum-likelihood estimation produces valid estimates without imputation as long as the MAR assumption holds. This is a key advantage over an analysis based on pre–post difference scores, which would discard a participant’s entire period contribution whenever a single timepoint is missing.

#### EXPLORATORY AND POST-HOC ANALYSES

Beyond the analyses pre-specified in the SAP, additional analyses may be added as study experience accrues, given the exploratory transdiagnostic nature of this trial. Any deviations from the prespecified plan, and any post-hoc analyses, are clearly labelled as such in the results manuscript and time-stamped in the trial registry where applicable.

The detailed statistical analysis plan was finalised, dated, and signed prior to database lock and is held in the trial master file at UMCG; it is available from the corresponding author on reasonable request.

### Monitoring

#### DATA MONITORING COMMITTEE

Because the intervention is a home-delivered dietary pattern of established foods with negligible anticipated risk, no formal Data Safety Monitoring Board (DSMB) was installed. This decision was endorsed by the accredited Medical Research Ethics Committee (METC BeBo Assen). No interim analyses are planned for early stopping; therefore, no stopping guidelines are specified.

#### TRIAL MONITORING

Trial monitoring is performed by an independent monitor from the UMCG. Given the negligible-risk classification, monitoring is risk-based and consists of source-data verification of a sample of participant files, verification of informed consent, and review of key study procedures. Monitoring is performed annually during the active conduct phase of the trial, with an end-of-study monitoring visit after database lock. All associated investigators are trained in Good Clinical Practice (GCP) and study procedures before contributing to the trial.

### Harms

All adverse events (AEs) reported spontaneously by participants or observed by investigators are recorded systematically. Adverse events are defined as any undesirable experience occurring to a subject during the study, whether or not considered related to the investigational product.

A serious adverse event (SAE) is any untoward medical occurrence or effect that:

- results in death;
- is life threatening (at the time of the event);
- requires hospitalisation or prolongation of existing inpatients’ hospitalisation;
- results in persistent or significant disability or incapacity;
- is a congenital anomaly or birth defect; or
- is any other important medical event that did not result in any of the outcomes listed above due to medical or surgical intervention but could have been based upon appropriate judgement by the investigator.

An elective hospital admission is not considered an SAE. Serious adverse events are reported by the sponsor through the ToetsingOnline web portal to the accredited METC within 7 days of first knowledge for SAEs that result in death or are life threatening (followed by a maximum of 8 days to complete the initial preliminary report); all other SAEs are reported within a maximum of 15 days of first knowledge.

In participants with bipolar disorder or schizophrenia spectrum disorder, psychiatric relapse and associated hospitalisation are common features of the underlying condition and are not anticipated to be related to the dietary intervention. If a participant in these diagnostic groups experiences a psychiatric relapse or hospitalisation during the intervention period, they are considered to have dropped out of the intervention; the event itself is recorded and reported in line with the SAE procedures described above.

### Ethics and dissemination

#### RESEARCH ETHICS APPROVAL

The trial received favourable ethical opinion from the accredited Medical Research Ethics Committee BeBo Assen (METC Stichting BEBO; registration number NL78755.056.21) on 11 January 2022 and was accepted for local execution by the Medical Ethics Committee of the University Medical Center Groningen. The trial was prospectively registered in the Dutch Overview of Medical-Scientific Research in the Netherlands (OMON: NL-OMON52339) before recruitment began.

#### CONSENT

After a potential participant is referred by their treating physician, a researcher contacts them by telephone, provides further information about the study, and sends a written information letter. Potential participants then have up to two weeks to consider participation. Participants who decide to take part receive their first home visit from a researcher, at which the informed consent form is read through and signed in the presence of the researcher before any study measurements are performed.

#### CONFIDENTIALITY

Personal information about potential and enrolled participants is collected, processed, and stored in accordance with the EU General Data Protection Regulation (GDPR) and the Dutch UAVG. Direct identifiers are stored separately from study data; the link is held by the research team. Study data are accessible only to authorised study personnel, the accredited METC, and the Dutch Health and Youth Care Inspectorate. Published data are fully de-identified.

#### ANCILLARY AND POST-TRIAL CARE

The sponsor (UMCG) holds insurance covering harm to participants resulting from study participation, in accordance with the Dutch Medical Research Involving Human Subjects Act (WMO).

After completion of the trial, no continued provision of the BrAIN food products is offered. To support participants who wish to continue the dietary pattern on their own, however, all participants retain the educational materials they received during the intervention period — including the BrAIN workbook, recipes, and shopping lists. Participants are not routinely informed of their individual results; aggregated trial results are disseminated via peer-reviewed publications and the public trial registry, in line with the dissemination policy described below.

#### PROTOCOL AMENDMENTS

Since the original favourable opinion of METC Stichting BEBO (NL78755.056.21), a total of seven amendments have been submitted: three substantial and four non-substantial. All substantial amendments were reviewed and approved by the METC before implementation; non-substantial amendments were recorded and filed by the sponsor. Each substantial amendment was communicated to the METC and - where applicable - to participants through an updated informed consent form, and was reflected in the trial registry where applicable.

The amendments to date are:

- **Non-substantial amendment 1 (January 2022):** professional redesign of recruitment materials (brochure and poster); no impact on study procedures or participant burden.
- **Substantial amendment 1 (October 2022):** addition of two new measurement domains—oral health (OHIP, oral health questionnaire developed by OZ Pruntel, dental visit, intra-oral photograph, oral swab) and stress/resilience (BRS, Brugha List of Threatening Experiences, CTQ, PSS-10, and HRV via ECG); accompanied by updated informed consent form (v5) and ABR form. Rationale: these domains address known transdiagnostic factors and were felt by the team to be feasibly integrated.
- **Non-substantial amendment 2 (January 2023):** administrative change—following accidental termination of approximately 50 mobile subscriptions during a UMCG system clean-up, the study mobile phone number became inactive and could not be reactivated. A new SIM was issued and the ICF was updated to v5.2.
- **Substantial amendment 2 (October 2023):** addition of the Mandometer device—a digital scale used to objectively measure the pattern and speed of food intake—accompanied by an updated research protocol (v4.1) and ICF (v5.3). Rationale: emerging evidence linking eating behaviour to gut microbiome and antipsychotic medication justified this additional pilot assessment.
- **Substantial amendment 3 (February 2024):** two modifications: (i) the ICF was updated to include a yes/no checkbox for an optional additional eating-pattern test; and (ii) the Mandometer assessment was relocated from V1 and V4 to V3 and V6 to reduce participant burden during the larger study visits. Accompanied by updated research protocol (v4.2) and ICF (v5.4).
- **Non-substantial amendment 3 (October 2024):** Administrative correction; the protocol footer date for amendment 5 had been omitted and was reinstated. In response to a routine clean-up of the ToetsingOnline portal, the projected end date of the study was updated to 2025 in the ABR form.
- **Non-substantial amendment 4 (April 2026):** formalisation of the realised diagnostic distribution. For patients living with Alzheimer’s disease (AD), the 15-month total study duration discouraged most from enrolling. As a result, only one AD participant was included, against 49 participants with Parkinson’s disease, 29 with bipolar disorder, and 28 with schizophrenia spectrum disorder. This amendment formalised the realised distribution and was approved by METC Stichting BEBO as a non-substantial amendment, given that it reflected actual recruitment patterns rather than a change in study design or methodology.

#### NON-PROTOCOL REFINEMENTS DOCUMENTED IN THE STATISTICAL ANALYSIS PLAN AND IN STANDARD OPERATING PROCEDURES

In addition to the formal protocol amendments listed above, five methodological refinements were made during trial conduct and were documented either in the statistical analysis plan (SAP) or as a Standard Operating Procedure (SOP) update, rather than through formal protocol amendments.

*Refinement of dietary adherence instrument.* The original protocol referenced the Dutch Healthy Diet Food Frequency Questionnaire (DHD-FFQ) for dietary adherence assessment. At the time of protocol registration, the disease-relevant MIND-NL-Eetscore-FFQ was under development through a collaboration with Wageningen University & Research and was operationally available, but its validation reference (Beers et al.) had not yet been published. The MIND-NL-Eetscore-FFQ was therefore used from study start as the dietary adherence instrument and was specified as such in the SAP.

*Refinement of primary analysis.* The primary analysis was refined from a linear mixed model on OQ-45 difference scores, as described in the original protocol and the public trial registry entry (NL-OMON52339), to a linear mixed-effects model with timepoint × treatment interaction. This refinement was made for methodological reasons: the timepoint × treatment approach handles missing observations more robustly under the missing-at-random assumption, makes more efficient use of partial data, and provides a more interpretable estimate of the differential change between conditions in a two-period crossover design.

*Addition of visual analogue scales for participant experience and tolerability.* Shortly after the start of recruitment in 2022, three visual analogue scales (VAS, 1–10) were added to the weekly intervention monitoring procedures: a VAS for gastrointestinal tolerability (also serving as an early-warning measure complementing routine adverse-event reporting), a VAS for recipe satisfaction, and a VAS for overall satisfaction with the dietary programme. These scales were administered at weeks 3, 6 and 9 of the BrAIN intervention period. Because they were process and feasibility measures rather than primary or secondary clinical outcomes, the change was implemented as a Standard Operating Procedure update by the trial team and was not submitted as a formal METC amendment. From the point of introduction onwards, the procedure was applied uniformly to all participants; the earliest few participants therefore did not complete the full set of VAS measurements. Data from these scales are reported in the diagnosis-focused outcome manuscripts.

*Refinement of fatigue instrument.* The original protocol specified the Short Fatigue Questionnaire (SFQ) for assessment of fatigue. During trial start-up the Fatigue Severity Scale (FSS) was selected instead, on the basis that it is more widely used internationally, has stronger established psychometric properties in both psychiatric and neurodegenerative populations, and allows direct comparison of findings with the broader literature. The change in instrument was implemented prior to enrolment of the first participant and was applied uniformly across the cohort; it was specified as such in the SAP. The two instruments are comparable in length and respondent burden, and both yield a unidimensional fatigue score, so the refinement did not affect participant procedures or burden.

*Refinement of microbiome sequencing platform.* The original protocol specified 16S rRNA gene sequencing for faecal microbiome analysis. Prior to sample analysis, this was updated to shotgun metagenomic sequencing, consistent with the protocol’s provision allowing methodological refinement when better alternatives become available. Shotgun metagenomic sequencing provides higher taxonomic resolution (species- and strain-level rather than genus-level) and direct functional gene profiling of microbial metabolic pathways, supporting more informative analyses of the inflammatory and metabolic pathways central to this trial. Faecal samples had already been collected and stored in line with the original procedure; no participant-facing procedures were affected by this change. The refinement was specified in the SAP prior to database lock.

The four SAP-related refinements were finalised, dated, and signed in the SAP prior to database lock; the SOP update was filed in the trial master file. The public registry entry has not been updated to reflect these refinements.

#### DISSEMINATION POLICY

The funding partner Hersenstichting (Brain Foundation Netherlands) is a non-profit organisation. The trial team is committed to transparent and unrestricted dissemination of all results, both positive and negative. Primary results are submitted for publication in peer-reviewed scientific journals and presented at relevant scientific conferences. A summary of results will be made available to participants, patient organisations, and the public in lay-language formats. Authorship follows the International Committee of Medical Journal Editors (ICMJE) criteria. The full trial protocol and de-identified individual participant data are available from the corresponding author upon reasonable request, after primary publication and in accordance with applicable data protection regulations.

#### Roles and Responsibilities

**Sponsor and Principal Investigator:** Prof. Dr. Iris E.C. Sommer, Center for Clinical Neuroscience and Cognition (CNC), University of Groningen, University Medical Center Groningen, P.O. Box 196, Internal Zip Code FA32, 9700 AD Groningen, The Netherlands. The sponsor is responsible for overall scientific oversight, study conduct, and reporting.

**Coordinating investigators:** Coordinating investigators: Sophie M. van Zonneveld, MSc and Greetje Huisman, MSc; both at the Center for Clinical Neuroscience and Cognition, University of Groningen, University Medical Center Groningen. They are responsible for day-to-day trial conduct, data collection, and data management, under supervision of the principal investigator and senior co-investigators. No formal steering committee or endpoint adjudication committee was instituted, given the nature of the intervention and the outcomes.

**Funder:** Hersenstichting (Brain Foundation Netherlands), grant ID J-06. The funder had no role in the design of the study, in data collection, analysis, or interpretation, in the writing of this manuscript, or in the decision to submit for publication.

## RESULTS

Funding was awarded by Hersenstichting in October 2020. Ethical approval was obtained from the accredited Medical Research Ethics Committee BeBo Assen on 11 January 2022, and the trial was prospectively registered at OMON (NL-OMON52339) before recruitment began. Recruitment commenced in February 2022; the first participant was enrolled on 7 March 2022, and the last on 6 May 2024. Follow-up was completed on 5 September 2025, marking the end of data collection. A total of 107 participants were enrolled across the four diagnostic groups: 49 PD, 29 BD, 28 SSD, and 1 AD

Data-analyses are currently ongoing. The primary results are reported in two separate manuscripts rather than a single transdiagnostic paper. Inspection of baseline characteristics revealed substantial heterogeneity between the Parkinson’s disease group and the psychiatric groups—including a mean age difference of approximately 18 years and divergent symptom profiles—which would preclude a coherent transdiagnostic narrative. This decision was based on baseline data only; no transdiagnostic outcome analysis was performed beforehand. Results are reported in a manuscript focusing on Parkinson’s disease and a manuscript focusing on the psychiatric groups (BD and SSD), both scheduled for submission in summer 2026.

## DISCUSSION

### Principal considerations

This protocol paper describes a randomised controlled crossover trial designed to evaluate whether an anti-inflammatory dietary pattern, termed the BrAIN diet, can improve global functioning and a broad set of clinical, cognitive, inflammatory, and gut-health outcomes in patients with bipolar disorder, schizophrenia spectrum disorder, Parkinson’s disease, and Alzheimer’s disease. The central rationale is that these four brain disorders, despite differing aetiologies, share two modifiable biological substrates, chronic low-grade neuroinflammation and gut-microbiome alterations, that are jointly addressable through diet. Existing dietary intervention trials in these populations have been largely confined to single-diagnosis studies of Mediterranean-style diets [43–44, 46], typically without parallel deep biomarker phenotyping. The present trial therefore combines an integrated anti-inflammatory dietary pattern with structured home delivery, weekly professional support, and integrated assessment of inflammatory, microbiome, gut-permeability, cognitive, and disease-specific outcomes — under a single shared protocol.

### Strengths

Several design features support the present approach. First, the two-period crossover design maximises statistical power while ensuring that all participants ultimately receive the intervention, with carryover and period effects formally tested in the analysis. Second, recruiting four brain-disorder populations under a single shared protocol - using identical assessments, sample handling, and analytical pipelines - allows mechanistic outcomes (inflammation, gut microbiome, gut-permeability biomarkers) to be examined both within and across diagnoses, supporting an integrated understanding of shared and disease-specific pathways. Third, the intervention applies an integrated anti-inflammatory dietary pattern [38] in a combined patient cohort and delivery format that have not previously been studied together. Fourth, structured home delivery with weekly dietitian contact addresses practical barriers to dietary change in these populations and is itself a novel implementation model. Fifth, the assessment battery is broad and mechanism-oriented, integrating inflammatory markers, microbiome composition, gut-permeability biomarkers, cognition, and disease-specific clinical scales; it was further expanded through approved amendments to include oral health, stress and resilience, and eating-behaviour measures. Finally, feasibility and acceptability are formally evaluated alongside clinical outcomes, using weekly visual analogue scales for gastrointestinal tolerability, recipe satisfaction, and overall programme satisfaction, so that implementation experience can be reported alongside efficacy.

### Limitations

Several limitations should be acknowledged, most of which reflect deliberate trade-offs in study design rather than oversights. First, the realised diagnostic distribution differs substantially from the original plan. Of 107 enrolled participants, only one had Alzheimer’s disease, against 49 with Parkinson’s disease, 29 with bipolar disorder, and 28 with schizophrenia spectrum disorder. As detailed under Protocol Amendments, the 15-month total study duration proved too demanding for AD-patients to enrol. Rather than pool these heterogeneous groups into a single underpowered analysis, primary clinical outcomes will be reported in two diagnosis-focused manuscripts (Parkinson’s disease; psychiatric disorders combined), with mechanistic outcomes additionally analysed across diagnoses. Per-diagnosis sample sizes still limit statistical power for individual disease effects. Second, the trial is open-label by necessity: participants prepare and consume meals themselves, which makes blinding to dietary condition unfeasible — a constraint shared by virtually all whole-diet intervention trials. To mitigate this, the primary outcome (OQ-45) is participant-reported and the most informative biomarker outcomes (microbiome composition, inflammatory markers, gut-permeability biomarkers) are objective and processed blinded to allocation in the laboratory. Third, the 24-week washout period may not fully eliminate carryover effects in all participants. This duration was chosen as a reasonable balance between minimising carryover and keeping total participation manageable; carryover and period effects are formally tested in the analysis, with sensitivity analyses where carryover is detected. Fourth, generalisability is bounded by the inclusion criteria. Participation requires adequate health literacy, household support, and cooking facilities, preconditions that follow directly from the intervention itself, but that exclude patients living in highly dependent care settings. The trial therefore informs feasibility and effect estimates for community-dwelling outpatients, which represent the population most likely to benefit from a home-delivered dietary programme in routine care. Finally, dietary adherence depends on sustained participant motivation, even with structured home delivery and weekly dietitian contact. This reflects a real-world condition rather than a study artefact: any dietary intervention translated to clinical practice will face the same constraint, and the trial’s adherence data will inform how implementable such an intervention is outside research conditions.

### Comparison with prior work

Set against this background, the present trial occupies a distinct position in the existing literature. It is the first prospective dietary intervention trial to recruit four brain-disorder populations under a single shared protocol, with harmonised assessments and a unified analytical framework. Previous dietary intervention studies in these populations have generally been diagnosis-specific and have provided limited integration of clinical and mechanistic outcomes [43–44, 46]. The present trial extends previous work by integrating clinical, cognitive, inflammatory, microbiome, and gut-permeability outcomes within a single protocol and by applying a randomised crossover design in which participants serve as their own controls. Together, these features provide a more comprehensive evaluation of dietary intervention effects across neuropsychiatric and neurodegenerative disorders.

### Clinical implications

If the trial confirms its hypothesised effects, several clinical implications follow. At the level of individual patient care, an anti-inflammatory dietary pattern could become a structured adjunct to standard pharmacological and psychosocial care, complementing rather than replacing existing treatments, particularly relevant in populations where overlapping symptoms such as cognitive dysfunction, fatigue, and low mood remain inadequately addressed by current therapies. At the level of clinical reasoning across diagnoses, a positive signal would support the broader development of *transdiagnostic* nutritional care, in which dietary intervention is targeted at shared inflammatory and gut–microbiome pathways rather than at single diagnostic categories - a framing that is increasingly discussed in both psychiatric and neurological care but is not yet reflected in clinical guidelines. At the level of service delivery, the home-delivery and weekly-support model evaluated here may serve as a template for how dietary interventions can be made feasible in-patient groups for whom self-directed dietary change is known to be difficult, informing the design of future implementation studies and nutrition services. At the same time, any such implications depend on the magnitude, durability, and population-specificity of the effects observed, which are addressed in the diagnosis-focused outcome manuscripts.

## Conclusions

This protocol paper formalises the design, conduct, and analytical framework of the No Guts, No Glory trial: a randomised crossover study of an anti-inflammatory dietary pattern across four brain-disorder populations recruited under a single shared protocol. By documenting the trial’s pre-specified outcomes, statistical approach, and protocol amendments alongside the diagnosis-focused outcome manuscripts, it provides the methodological reference against which the trial’s clinical and mechanistic findings can be interpreted. In doing so, it provides a methodological foundation for future dietary-intervention research targeting shared gut–brain and inflammatory pathways across brain disorders.

## Data Availability

All data produced in the present study are available upon reasonable request to the authors

## Acknowledgements

The authors thank the participating patients and their families, the participating outpatient clinics, the dietitians, interns, study nurses, and laboratory personnel who contributed to the trial. We are grateful to the members of the patient advisory board for their input throughout the study, to Jesca de Jager (UMCG) for serving as independent randomisation officer, and to the patient associations for raising awareness of the study within their communities.

## Data availability

De-identified individual participant data underlying the published results, the data dictionary, and the analytic code will be made available from the corresponding author upon reasonable request. Access is subject to a data-sharing agreement and applicable data-protection regulations (GDPR/UAVG). Requests will be reviewed by the principal investigator; data will be shared for academic, non-commercial use.

## Authors’ contributions

IECS conceived the study and obtained funding. SMVZ developed the detailed protocol and intervention procedures. EJvdO designed the dietary intervention (BrAIN diet) and supervised the dietitians in training. JON provided statistical and methodological input on the trial design and analysis plan. BCMH supervised SMVZ and GH and contributed to the clinical design of the trial in the psychiatric groups. SMVZ wrote a substantial amendment to investigate oral health and the oral microbiome; GH wrote a substantial amendment to investigate stress resilience in the psychiatric groups; TAWS wrote a substantial amendment to investigate eating behaviour as a pilot study. SMVZ and GH performed data collection, supported by the wider study team. SMVZ drafted the manuscript, with TAWS contributing to drafting of specific sections. GH, EJvdO, JON, BCMH, and IECS critically revised the manuscript for important intellectual content. IECS supervises the overall project. All authors read and approved the final version of the manuscript and agree to be accountable for all aspects of the work.

## Abbreviations

AD: Alzheimer’s disease
AE: adverse event
AIDP: anti-inflammatory dietary pattern
BAC: Brief Assessment of Cognition
BD: bipolar disorder
BMI: body mass index
BPRS: Brief Psychiatric Rating Scale
BrAIN: Brain Anti-Inflammatory Nutrition
BRS: Brief Resilience Scale
CRP: C-reactive protein
CTQ-SF: Childhood Trauma Questionnaire-Short Form
DaU: diet-as-usual
DHD-FFQ: Dutch Healthy Diet-Food Frequency Questionnaire
DII: Dietary Inflammatory Index
DSMB: Data Safety Monitoring Board
ECG: electrocardiography
eCRF: electronic case record form
EQ-5D: EuroQoL 5 dimensions
FDR: false discovery rate
FFQ: Food Frequency Questionnaire
FSS: Fatigue Severity Scale
GAF: Global Assessment of Functioning
GCP: Good Clinical Practice
GDPR: General Data Protection Regulation
GSRS: Gastrointestinal Symptom Rating Scale
HDL: high-density lipoprotein
HRV: heart rate variability
IADL: Instrumental Activities of Daily Living
ICF: informed consent form
ICMJE: International Committee of Medical Journal Editors
I-FABP: intestinal fatty acid-binding protein
IL: interleukin
I.ROC: Individual Recovery Outcomes Counter
IRRID: International Registered Report Identifier
LBP: lipopolysaccharide-binding protein
MDS-UPDRS: Movement Disorders Society Unified Parkinson Disease Rating Scale
METC: Medical Research Ethics Committee
MIND: Mediterranean-DASH Intervention for Neurodegenerative Delay
NGNG: No Guts, No Glory
NMSQ: Non-Motor Symptoms Questionnaire
OHIP: Oral Health Impact Profile
OMON: Overview of Medical-Scientific Research in the Netherlands
OQ-45: Outcome Questionnaire-45
PD: Parkinson’s disease
PSS-10: Perceived Stress Scale-10
SAE: serious adverse event
SAP: Statistical Analysis Plan
sCD14: soluble CD14
SCFA: short-chain fatty acids
SPIRIT: Standard Protocol Items: Recommendations for Interventional Trials
SSD: schizophrenia spectrum disorders
TNF: tumour necrosis factor
UAVG: Dutch Implementing Act for the GDPR
UMCG: University Medical Center Groningen
VAS: visual analogue scale
WMO: Dutch Medical Research Involving Human Subjects Act.

## Supplementary

**Supplementary Table S1.** **SPIRIT 2013 Checklist** This trial protocol is reported in accordance with the SPIRIT 2013 Statement (Standard Protocol Items: Recommendations for Interventional Trials), which was the version in effect at the time of protocol development in 2021. For each of the 33 SPIRIT 2013 checklist items, the table below summarises how the item is addressed in the manuscript and indicates the corresponding section. Items 1–5 fall under Administrative information; items 6–8 under Introduction; items 9–22 under Methods; items 24–31 under Ethics and dissemination; and items 32–33 under Appendices.

| Item | Item No | Description | Addressed in protocol (summary) | Section |
| --- | --- | --- | --- | --- |
| ADMINISTRATIVE INFORMATION |  |  |  |  |
| Title | 1 | Descriptive title identifying the study design, population, interventions, and, if applicable, trial acronym. | Title page identifies design (randomised controlled crossover trial), population (psychiatric and neurodegenerative disorders), intervention (anti-inflammatory dietary pattern), and acronym (NGNG / BrAIN diet). | Title page |
| Trial registration | 2a | Trial identifier and registry name. If not yet registered, name of intended registry. | OMON NL-OMON52339 reported in Abstract (Trial Registration), Methods (Study design) and Ethics and dissemination (Research ethics approval). | Abstract; Methods – Study design; Ethics – Research ethics approval |
|  | 2b | All items from the WHO Trial Registration Data Set. | WHO Trial Registration Data Set items reported across the public registry entry (NL-OMON52339) and the manuscript: title, registration date, sponsor, funder, contact, conditions studied, intervention, eligibility, design, sample size, primary and secondary outcomes. | Title page; Abstract; Methods – Study design, Eligibility, Outcomes; Roles and Responsibilities |
| Protocol version | 3 | Date and version identifier. | Manuscript date in header. Approved protocol version 4.2 (February 2024) referenced under Protocol amendments. | Title page; Ethics – Protocol amendments |
| Funding | 4 | Sources and types of financial, material, and other support. | Funding section: Hersenstichting (Brain Foundation Netherlands), grant ID J-06; funder role described under Roles and Responsibilities. | Funding; Roles and Responsibilities |
| Roles and responsibilities | 5a | Names, affiliations, and roles of protocol contributors. | Authors and affiliations on title page; Authors' contributions section. | Title page; Authors' contributions |
|  | 5b | Name and contact information for the trial sponsor. | Sponsor: UMCG, with PI Prof. Dr. Iris E.C. Sommer; full address and email under Roles and Responsibilities. | Roles and Responsibilities |
|  | 5c | Role of sponsor and funders in design, conduct, analysis, reporting, and decision to submit. | Funder role explicitly stated under Roles and Responsibilities and Funding: 'The funder had no role in the design of the study, in data collection, analysis, or interpretation, in the writing of this manuscript, or in the decision to submit for publication.' | Roles and Responsibilities; Funding |
|  | 5d | Composition, roles, and responsibilities of the coordinating center, steering committee, endpoint adjudication committee, data management team, and other groups overseeing the trial. | Coordinating investigators (SMVZ, GH) and sponsor/PI (IECS) described under Roles and Responsibilities. Explicitly stated: no formal steering committee or endpoint adjudication committee was instituted given the nature of the intervention. Independent monitor described under Trial monitoring; data management team described under Data management. | Roles and Responsibilities; Methods – Data management; Monitoring – Trial monitoring |
| INTRODUCTION |  |  |  |  |
| Background and rationale | 6a | Description of research question and justification, including summary of relevant studies (published and unpublished) examining benefits and harms for each intervention. | Introduction sections: Background and rationale, Neuroinflammation as a shared pathway, The gut-brain axis in brain disorders, Diet as modifier of inflammation and the microbiome. Prior dietary intervention studies (Mediterranean, MIND, Shivappa DII) summarised. | Introduction |
|  | 6b | Explanation for choice of comparators. | Comparator is diet-as-usual (no intervention). Rationale: open-label nutritional trial, no placebo diet feasible; crossover within-subject control. Described under Study design and Discussion (Limitations). | Methods – Study design; Discussion – Limitations |
| Objectives | 7 | Specific objectives or hypotheses. | Primary and secondary hypotheses listed in Introduction (Objectives and hypotheses). | Introduction – Objectives and hypotheses |
| Trial design | 8 | Description of trial design, including type of trial (e.g. parallel group, crossover, factorial, single group), allocation ratio, and framework (e.g. superiority, equivalence, non-inferiority, exploratory). | Open-label, randomised controlled, two-period crossover trial; 12-week intervention periods, 24-week washout; 1:3 allocation; exploratory framework. Described under Methods – Study design. | Methods – Study design |
| METHODS: PARTICIPANTS, INTERVENTIONS AND OUTCOMES |  |  |  |  |
| Study setting | 9 | Description of study settings (e.g. community clinic, academic hospital) and list of countries where data will be collected. Reference to where list of study sites can be obtained. | Outpatient clinics in northern Netherlands (UMCG, Martini Hospital, care centre Maartenshof, Parkinson Expertise Centers Groningen and Leeuwarden, MCL, GGZ Lentis, GGZ Friesland). All visits at participants' homes or at UMCG. Country: Netherlands. Described under Trial Setting. | Methods – Trial Setting |
| Eligibility criteria | 10 | Inclusion and exclusion criteria for participants. If applicable, eligibility criteria for study centers and individuals who will perform the interventions. | Inclusion (clinical diagnosis BD/SSD/PD/AD; living in Drenthe/Friesland/Groningen; cognitive capacity; 18–80 yr; Dutch language; able to follow diet and home visits; living independently) and exclusion criteria (pregnancy; severe under/overweight; severe bowel/liver disease/pancreatitis; incompatible diet/allergies; already AIDP — MIND-NL-Eetscore $\geq 10$; antibiotic/probiotic use) listed under Eligibility criteria. | Methods – Eligibility criteria |
| Interventions | 11a | Interventions for each group with sufficient detail to allow replication, including how and when they will be administered. | BrAIN diet described in detail under Intervention (BrAIN diet, Intervention delivery). Components: | Methods – Intervention; |
|  |  |  | Shivappa DII, MIND, Dutch Healthy Diet guidelines. Delivered via weekly home-delivered food boxes, weekly dietitian counselling (home or video), recipes, instructional videos. Detailed food choice list in Supplementary Table S3; example menu in Supplementary Table S4; recipes publicly available [50]. | Supplementary Appendices S3–S4 |
|  | 11b | Criteria for discontinuing or modifying allocated interventions for a given trial participant. | Modification/discontinuation criteria (adverse event, participant request, change in clinical status) described under Intervention modifications and discontinuation criteria. Adaptation of food list preferred over discontinuation. | Methods – Intervention modifications and discontinuation criteria |
| | 11c | Strategies to improve adherence to intervention protocols, and any procedures for monitoring adherence. | Adherence supported through home delivery, individualised meal plan, weekly dietitian contact, instructional videos. Monitored via MIND-NL-Eetscore-FFQ ($\geq 10$ threshold), 5-day food diary, FFQ, 24-hour recall, dietary coach 80% rating, Goldberg method for energy plausibility. Described under Adherence monitoring. | Methods – Adherence monitoring |
|  | 11d | Relevant concomitant care and interventions that are permitted or prohibited during the trial. | Standard medical care, disease-specific pharmacological treatment, allied-health care continue. Stable chronic medication requested; antibiotic/regular probiotic use is exclusion criterion at baseline; new dietary patterns/supplementation discouraged. Described under Concomitant care. | Methods – Concomitant care |
| Outcomes | 12 | Primary, secondary, and other outcomes, including specific measurement variable, analysis metric, method of aggregation, and time point. Explanation of clinical relevance recommended. | Primary: OQ-45 (global functioning); analysis metric is the timepoint × treatment interaction in a linear mixed-effects model comparing the BRAIN diet with the control condition (diet-as-usual; DaU); assessed at V1–V6 (weeks 0/12/24/36/48/60). Secondary outcomes (cognition, EQ-5D, FSS, GSRS, Bristol Stool Chart, BMI/metabolic, gut permeability biomarkers, microbiome, inflammation/oxidative stress, disease-specific scales) listed under Outcome measures. Additional outcomes (oral health, stress/resilience, eating behaviour) added through amendments. | Methods – Outcome measures; Table 1 |
| Participant timeline | 13 | Time schedule of enrolment, interventions (including any run-ins and washouts), assessments, and visits for participants. A schematic diagram is highly recommended. | Schematic timeline (Figure 1) and SPIRIT Schedule of Enrolment, Interventions and Assessments (Table 1). Schedule of assessments per visit detailed in Supplementary Table S2. | Methods – Study design; Figure 1 & Table 1; Supplementary Table S2 |
| Sample size | 14 | Estimated number of participants needed to achieve study objectives and how it was determined, including clinical and statistical assumptions. | Cohen's $d = 0.5$; $\alpha = 0.05$ (two-sided); $\beta = 0.20$ (80% power); $n = 64$ required for primary analysis; planned 100 participants accounting for 30% dropout. Described under Sample size. | Methods – Sample size |
| Recruitment | 15 | Strategies for achieving adequate participant enrollment to reach target sample size. | Referrals through outpatient clinics, patient organisations, Hersenonderzoek.nl registry, presentations at patient associations, newsletters, repeat invitations, targeted outreach. Home visits and travel reimbursement to lower barriers. Described under Recruitment. | Methods – Recruitment |
| METHODS: ASSIGNMENT OF INTERVENTIONS |  |  |  |  |
| Allocation – Sequence generation | 16a | Method of generating the allocation sequence and list of any factors for stratification. Details of any planned restriction (e.g. blocking) should be in a separate document unavailable to those who enrol participants or assign interventions. | Sequence generated using Sealed Envelope (UK); 1:3 allocation; permuted block randomisation, block size 4; stratification by diagnosis and sex. Per-stratum CSV files held by independent investigator separately from the recruitment team. | Methods – Randomisation: Sequence generation |
| Allocation concealment mechanism | 16b | Mechanism of implementing the allocation sequence (e.g. central telephone; sequentially numbered, opaque, sealed envelopes), describing any steps to conceal the sequence until interventions are assigned. | Pre-generated lists held by independent investigator. Recruiting researcher provided diagnosis and sex only; received next allocation plus sequential study ID (NG001...). Lists transferred to study team only after recruitment closed. | Methods – Randomisation: Allocation concealment |
| Implementation | 16c | Who will generate the allocation sequence, who will enrol participants, and who will assign participants to interventions. | Sequence generation, allocation, and assignment by independent investigator (Jesca de Jager, UMCG). Recruiting researchers screened, consented, and enrolled but had no access to randomisation lists. | Methods – Randomisation: Implementation; Acknowledgements |
| Blinding (masking) | 17a | Who will be blinded after assignment to interventions (e.g. trial participants, care providers, outcome assessors, data analysts), and how. | Open-label: participants and intervention providers not blinded (dietary intervention with weekly counselling). Outcome assessors not formally blinded; primary outcome (OQ-45) is participant-reported. Laboratory personnel blinded to allocation (samples coded NG001...); statisticians not blinded. | Methods – Randomisation: Blinding |
|  | 17b | If blinded, circumstances under which unblinding is permissible, and procedure for revealing a participant's allocated intervention during the trial. | Not applicable – open-label design; no formal unblinding procedure required. | Methods – Randomisation: Blinding |
| METHODS: DATA COLLECTION, MANAGEMENT, AND ANALYSIS |  |  |  |  |
| Data collection methods | 18a | Plans for assessment and collection of outcome, baseline, and other trial data, including any related processes to promote data quality and a description of study instruments along with their reliability and validity, if known. Reference to where data collection forms can be found, if not in the protocol. | Self-report on paper/tablet at home; cognitive tasks by trained researchers; blood by trained personnel/mobile phlebotomy; faecal samples self-collected via standardised kit. All instruments are validated and listed in Table 2 with language version and validation references. GCP training for all assessors. | Methods – Data collection methods; Table 1 |
|  | 18b | Plans to promote participant retention and complete follow-up, including list of any outcome data to be collected for participants who discontinue or deviate from intervention protocols. | Retention strategies (weekly dietitian contact, scheduling flexibility, travel reimbursement) under Retention. Subsequent assessments still attempted for participants who discontinue, in line with intention-to-treat, where consent permits. | Methods – Retention |
| Data management | 19 | Plans for data entry, coding, security, and storage, including any related processes to promote data quality. Reference to where details of data management procedures can be found, if not in the protocol. | REDCap (v16.0.17) hosted by UMCG IT Services; role-based access; encryption in transit; audit trail; coded study identifier (NG001...) with key held separately; field-level validation, range checks, branching logic; verification by trained interns; periodic Data Quality module runs; source data verification by independent monitor; GCP training. Records retained 15 years; biological samples 5 years. | Methods – Data management |
| Statistical methods | 20a | Statistical methods for analysing primary and secondary outcomes. Reference to where other details of the statistical analysis plan can be found, if not in the protocol. | Primary analysis: linear mixed-effects model on OQ-45, with timepoint × treatment interaction, period and sequence as fixed effects, participant as random intercept, age and sex covariates. Secondary outcomes: same model structure per outcome; Benjamini–Hochberg FDR correction; Cohen's d. Detailed SAP held in trial master file at UMCG and available on reasonable request. | Methods – Statistical analysis: Primary analyses, Secondary analyses |
|  | 20b | Methods for any additional analyses (e.g. subgroup and adjusted analyses). | Pre-specified subgroup analyses: (i) psychiatric (BD, SSD) vs neurodegenerative (PD, AD); (ii) per-diagnosis (exploratory). Microbiome analyses: alpha/beta diversity, pre-specified taxa, hypothesis-driven genera. Carryover and period effects formally tested; sensitivity analyses where carryover detected. Exploratory and post-hoc analyses are clearly labelled. | Methods – Statistical analysis: Carryover and period effects, Secondary analyses, Microbiome analyses, Exploratory and post-hoc analyses |
|  | 20c | Definition of analysis population relating to protocol nonadherence (e.g. as-randomized analysis), and any statistical methods to handle missing data (e.g. multiple imputation). | Both intention-to-treat and per-protocol analyses pre-specified. Per-protocol excludes participants below MIND-NL-Eetscore <10 at end of intervention or <80% delivered-products consumed. Linear mixed-effects model handles missing data under MAR via maximum-likelihood estimation; no imputation required. | Methods – Adherence monitoring; Statistical analysis: Primary analyses, Missing data |
| METHODS: MONITORING |  |  |  |  |
| Data monitoring | 21a | Composition of the Data Monitoring Committee (DMC); summary of its role and reporting structure; statement of whether it is independent from the sponsor and competing interests; and reference to where further details about its charter can be found, if not in the protocol. Alternatively, an explanation of why a DMC is not needed. | No formal DSMB installed; explained under Monitoring – Data monitoring committee. Decision endorsed by METC BeBo Assen given negligible-risk classification (home-delivered dietary pattern of established foods). | Monitoring – Data monitoring committee |
|  | 21b | Description of any interim analyses and stopping guidelines, including who will have access to these interim results and make the final decision to terminate the trial. | No interim analyses planned for early stopping; no stopping guidelines specified. Stated under Monitoring – Data monitoring committee. | Monitoring – Data monitoring committee |
| Harms | 22 | Plans for collecting, assessing, reporting, and managing solicited and spontaneously reported adverse events and other unintended effects of trial interventions or trial conduct. | AE and SAE definitions, reporting workflow (sponsor → ToetsingOnline → METC: 7 days for life-threatening/death, 15 days for other SAEs), and active monitoring at every visit and weekly dietitian contact (BrAIN period). VAS for GI tolerability at weeks 3, 6, 9. Psychiatric relapse in BD/SSD treated as dropout but reported as SAE per procedures. Described under Harms. | Ethics and dissemination – Harms |
| Auditing | 23 | Frequency and procedures for auditing trial conduct, if any, and whether the process will be independent from investigators and the sponsor. | Risk-based monitoring by independent monitor from UMCG: source-data verification of a sample of files, verification of informed consent, review of key procedures. Described under Trial monitoring. | Monitoring – Trial monitoring |
| ETHICS AND DISSEMINATION |  |  |  |  |
| Research ethics approval | 24 | Plans for seeking REC/IRB approval. | Favourable opinion of METC Stichting BEBO (NL78755.056.21) on 11 January 2022; locally accepted by METC UMCG. Described under Research ethics approval. | Ethics and dissemination – Research ethics approval |
| Protocol amendments | 25 | Plans for communicating important protocol modifications (e.g. changes to eligibility criteria, outcomes, analyses) to relevant parties (e.g. investigators, RECs/IRBs, trial participants, trial registries, journals, regulators). | Seven amendments to date (3 substantial, 4 non-substantial), each described under Protocol amendments with date, content, and review status. Substantial amendments reviewed and approved by METC before implementation; non-substantial filed by sponsor. Future modifications to be communicated to METC, regulators, participants where applicable, and the trial registry. Two non-amendment refinements documented in the SAP also disclosed. | Ethics and dissemination – Protocol amendments; Non-protocol refinements |
| Consent or assent | 26a | Who will obtain informed consent or assent from potential trial participants or authorized surrogates, and how. | Researcher contacts referred patient by phone; sends information letter; ≥2-week consideration period; written informed consent signed in researcher's presence at first home visit before any study procedures. Cognitive capacity reassessed at V3. Described under Consent. | Ethics and dissemination – Consent |
|  | 26b | Additional consent provisions for collection and use of participant data and biological specimens in ancillary studies, if applicable. | ICF v5.4 includes optional yes/no checkbox for additional eating-pattern test (substantial amendment 3). Biological samples retained 5 years for future research only with explicit additional consent (Data management). | Ethics – Consent; Methods – Data management |
| Confidentiality | 27 | How personal information about potential and enrolled participants will be collected, shared, and maintained in order to protect confidentiality before, during, and after the trial. | GDPR/UAVG-compliant; direct identifiers stored separately from study data; key held by research team; access restricted to authorised personnel, METC, Health and Youth Care Inspectorate; published data fully de-identified. Described under Confidentiality and Data management. | Ethics – Confidentiality; Methods – Data management |
| Declaration of interests | 28 | Financial and other competing interests for principal investigators for the overall trial and each study site. | Conflicts of interest section: 'The authors declare that they have no competing interests.' | Conflicts of interest |
| Access to data | 29 | Statement of who will have access to the final trial data set, and disclosure of contractual agreements that limit such access for investigators. | Investigators and authorised study personnel have access to the final trial data set; no contractual limits on access. De-identified IPD, data dictionary, and statistical code available from corresponding author on reasonable request after primary publication, subject to a data-sharing agreement. | Ethics – Dissemination policy; Data availability |
| Ancillary and post-trial care | 30 | Provisions, if any, for ancillary and post-trial care, and for compensation to those who suffer harm from trial participation. | UMCG holds insurance per WMO. After trial, no continued provision of BRAIN food products; participants retain workbook, recipes, shopping lists. Aggregated results disseminated; participants not routinely informed of individual results. Described under Ancillary and post-trial care. | Ethics – Ancillary and post-trial care |
| Dissemination policy | 31a | Plans for investigators and sponsor to communicate trial results to participants, health care professionals, the public, and other relevant groups (e.g. via publication, reporting in results databases, or other data-sharing arrangements), including any publication restrictions. | Hersenstichting is a non-profit funder; no publication restrictions. Results submitted to peer-reviewed journals, presented at conferences; lay-language summary for participants, patient organisations, and the public. Described under Dissemination policy. | Ethics – Dissemination policy |
|  | 31b | Authorship eligibility guidelines and any intended use of professional writers. | Authorship follows ICMJE criteria. No professional medical writers used. Stated under Dissemination policy. | Ethics – Dissemination policy |
|  | 31c | Plans, if any, for granting public access to the full protocol, participant-level data set, and statistical code. | Full protocol and de-identified IPD plus statistical code available from corresponding author upon reasonable request after primary publication; subject to data-sharing agreement and applicable data-protection regulations. | Ethics – Dissemination policy; Data availability |
| APPENDICES |  |  |  |  |
| Informed consent materials | 32 | Model consent form and other related documentation given to participants and authorized surrogates. | Informed consent form (ICF v5.4) and participant information letter held in the trial master file at UMCG and submitted to METC. Available from corresponding author on request. | Ethics – Consent (referenced) |
| Biological specimens | 33 | Plans for collection, laboratory evaluation, and storage of biological specimens for genetic or molecular analysis in the current trial and for future use in ancillary studies, if applicable. | Blood (gut permeability biomarkers: zonulin, LBP, sCD14, I-FABP, EndocAb; inflammation: hs-CRP, IL-6, TNF-α, free serum thiols; metabolic: triglycerides, HDL); faecal samples (shotgun metagenomic sequencing, faecal pH, SCFAs, redox potential, calprotectin); oral swab (oral microbiome, added through amendment). Self-collection kit for faeces. Samples retained 5 years; future research only with explicit additional consent. | Methods – Outcome measures, Data collection methods, Data management; Ethics – Protocol amendments |

**Supplementary Table S2.**
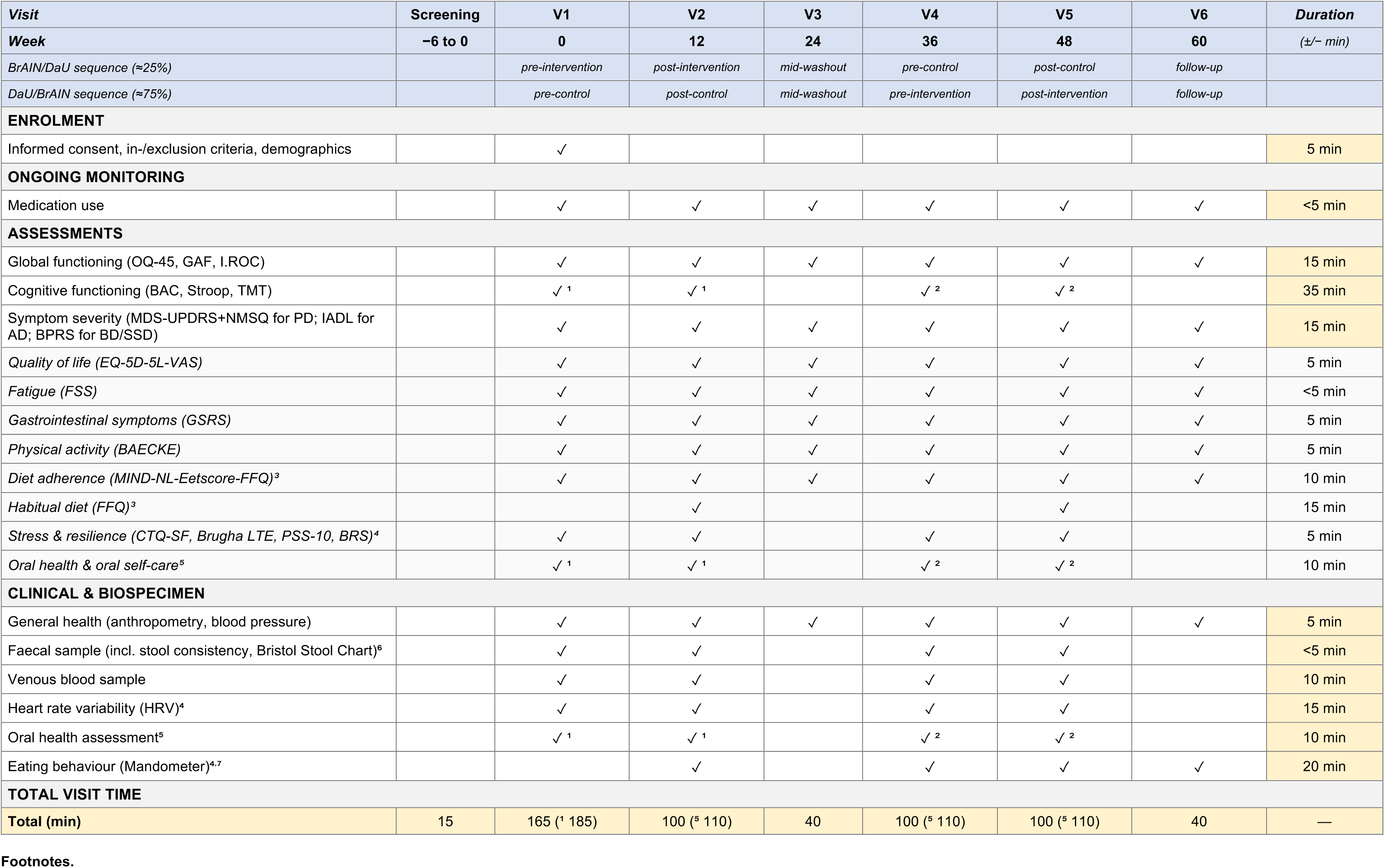
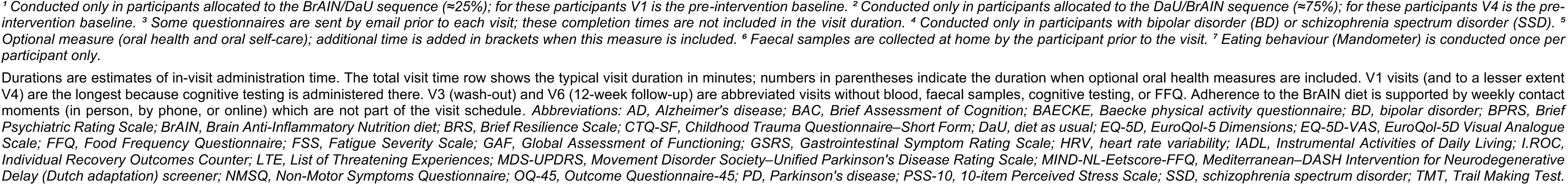
Schedule of assessments per visit, including approximate duration.

**Supplementary Table S3.** BrAIN dietary pattern – food choice list.

| Food group | Always a good choice | In moderation | Better to avoid |
| --- | --- | --- | --- |
| **Vegetables** | Unprocessed fresh and frozen vegetables<br>Particular attention:<br>• Kale (boerenkool)<br>• Endive (andijvie)<br>• Purslane (postelein)<br>• Spinach<br>• Lettuce<br>• Broccoli<br>• Cabbage<br>• Carrots<br>• Onions | Canned and dried vegetables without added salt or sugar<br>Sauerkraut (zuurkool) | Vegetables in cans or jars with added salt or sugar<br>Vegetables in cream sauce |
| **Fruit** | All unprocessed and frozen fruit (avocado is classified as fruit here)<br>Particular attention:<br>• Strawberries<br>• Berries<br>• Apples with peel<br>• Oranges | Puréed fruit<br>Dried fruit (unsweetened)<br>Canned fruit<br>Coconut | Fruit juice<br>Sweetened dried fruit |
| **Bread and breakfast cereals** | Wholegrain bread, optionally with seeds, kernels, nuts or dried fruit<br>Oatmeal<br>Muesli without added sugar (optionally with grains, nuts, dried fruit)<br>All Bran (plain) | Brown bread<br>Crispbread (knäckebröd) or crackers with fibre (>6 g/100 g)<br>Wholegrain rusks<br>Brinta (Dutch wholewheat porridge)<br>Bambix (Dutch infant wholegrain porridge)<br>Molenaar 7-grain breakfast | White bread, sweet bread, croissants, pancakes, wraps, or pizza bases made from white flour<br>Crispbread (knäckebröd), rusks (white)<br>Crunchy muesli<br>Breakfast cereals with added salt or sugar (e.g. Choco Pops, Smacks, Rice Crispies, Frosties)<br>Cornflakes<br>Drinkable breakfast products<br>Pizza rolls |
| **Pasta, noodles, rice** | Brown rice (zilervliesrijst), wholegrain pasta, wholegrain couscous, wholegrain bulgur, quinoa<br>Wholegrain noodles<br>Wholegrain pita / tortilla / wraps | Multigrain rice | White rice, white pasta, white couscous, white noodles<br>Plain pita / tortilla (white) |
| **Potatoes** | Boiled, steamed, or baked potatoes<br>Sweet potato<br>Pomtajer (Surinamese taro)<br>Yam | Pan-fried potatoes prepared with olive oil and without salt | Mashed potato (commercial)<br>Potato products with added salt (potato wedges, fries, rösti, potato croquettes) |
| **Fish** | All unprocessed fish (fresh, steamed, smoked, freshly frozen)<br>• Cod (kabeljauw)<br>• Haddock (schelvis)<br>• Tilapia, sole (tong), whiting (wijting)<br>• Scallops (coquilles), squid (inktvis), mussels, oysters<br>Unprocessed and not canned moderately fatty fish: shrimp, trout, plaice (schol), tuna, sea bass, wolffish (zeewolf), halibut, pangasius<br>Particular attention to oily fish:<br>• Salmon, smoked salmon, salmon trout<br>• Herring, mackerel, eel<br>• Sprat, anchovies, sardines | Soused herring (zure haring) | Fish products containing less than 70% fish<br>Breaded fish<br>Pan-fried fish (commercial)<br>Deep-fried fish<br>Salted fish<br>Canned fish |
| **Legumes** | Fresh legumes<br>Canned or jarred legumes with $\leq 200$ mg sodium per 100 g and no added sugar | — | Canned or jarred legumes with $> 200$ mg sodium per 100 g and/or added sugar |
| **Meat and meat products** | The following unprocessed meats:<br>• Chicken<br>• Turkey | Unprocessed red meat:<br>• Beef<br>• Pork | All processed meats, such as hamburgers, beef olive (rundervink), sausages, shoarma |
|  | • Duck<br>• Goose<br>• Rabbit<br>Slices of the above unprocessed meats may also be used as bread topping. | • Goat<br>• Lamb / mutton<br>• Horse meat | Breaded meat<br>Pre-seasoned ready-to-cook meat<br>All cured meat products (deli meats, ham, bacon, sausage varieties, etc.) |
| **Meat substitutes** | Required nutritional criteria (per 100 g):<br>• Saturated fat: <2.5 g<br>• Trans fat: <0.1 g<br>• Sodium: ≤450 mg<br>• No added sugar<br>• Iron: >0.8 mg<br>• Vitamin B12: ≥0.06 µg<br>• Vitamin B1: ≥0.06 mg<br>• Protein: >12 en%<br>Examples meeting these criteria:<br>• Tofu (tahoe)<br>• Tempeh<br>• Quorn<br>• Legumes<br>• Unsalted nuts<br>Plain organic tofu and tempeh products, mushroom-based mince products, and other minimally processed plant-based products meeting the criteria above. | — | Ready-to-eat meat substitutes that do not meet the nutritional criteria specified in the left column |
| **Eggs** | Boiled egg<br>Poached egg<br>Fried egg prepared with olive oil | — | — |
| **Nuts** | All unsalted, roasted or unroasted nuts<br>Nuts without additives (including peanuts, seeds and kernels)<br>Self-seasoned nuts | — | All salted and sugared nuts and peanuts<br>Coated savoury nuts (borrelnoten — all varieties) |
|  | Nut spread (100%)<br>Peanut butter (100%)<br>Particular attention:<br>• Walnuts<br>• Unsalted student-blend nuts (studentenhaver)<br>• Unsalted premium nut mix (elitehaver)<br>• Unsalted tropical mix |  | Tiger nuts (tiggernoten)<br>Japanese mix<br>Nuts coated with chocolate (e.g. M&M's)<br>Nut spread or peanut butter with added salt or sugar |
| **Milk and dairy products** | Buttermilk<br>Plain low-fat or semi-skimmed yoghurt<br>Plain low-fat or semi-skimmed quark (kwark)<br>Kefir<br>Plain plant-based protein drinks and desserts with added calcium and B12<br>Plain Turkish, Greek, or Bulgarian yoghurt (0% or 2%) | Low-fat and semi-skimmed milk<br>Fruit yoghurt without added sugar or preservatives<br>Plain Turkish, Greek, or Bulgarian yoghurt (5%)<br>Semi-skimmed coffee creamer | Whole-fat dairy products<br>Custard (vla), including cream-based custard<br>Pudding<br>Yoghurt drinks<br>Fruit yoghurt with added sugar<br>Plant-based protein drinks with >6 g sugar per 100 g<br>Soft-serve ice cream, water ice, dairy ice cream<br>Smoothies<br>Plain Turkish, Greek, or Bulgarian yoghurt (10%) or fruit-flavoured varieties<br>Coffee creamer (whole-fat)<br>Sour cream, whipping cream, cooking cream, crème fraîche |
| **Cheese** | 10+, 20+, and 30+ cheeses (plain, with cumin, or with herbs) — Dutch classification: ≤10/20/30% fat in dry matter<br>Soft goat's cheese<br>Mozzarella<br>Dairy spread without additives<br>Cottage cheese (huttenkäse) | Camembert<br>Brie<br>Aged cheese (oude kaas) | 48+ cheeses (plain, with cumin, or with herbs)<br>Smoked cheese<br>Boursin |
| **Spreads and cooking fats** | Olive oil<br>Low-fat margarine (halvarine) with added omega-3 fatty acids | Other plant-based oils<br>Liquid margarine and liquid frying fat (bottled)<br>Diet margarine<br>Diet low-fat margarine (dieethalvarine) | Coconut fat<br>Butter (roomboter)<br>Solid frying fat<br>Margarine and frying fat sold in wrappers (block form) |
| **Beverages** | Water<br>Green and black tea<br>Filter coffee without sugar | Kombucha<br>Yerba mate<br>Red wine<br>Berry juice without added sugar (e.g. Zonnatura) | Soft drinks (including light and zero versions)<br>All vegetable and fruit juices<br>Boiled coffee or coffee made with a cafetière (French press)<br>Lemonade or squash (including sugar-free varieties)<br>Energy drinks<br>Sports drinks |
| **Soup** | Home-made meal soup with $\leq 2$ g salt per portion, used as a replacement for the main hot meal | — | Stock / bouillon (commercial)<br>Ready-made soup<br>Instant cup soup (cup-a-soup) |
| **Sauces** | Home-made sauce without added salt or sugar | — | All ready-made sauces |
| **Herbs and spices** | All fresh herbs and spices<br>Freshly frozen herbs<br>Particular attention:<br>• Turmeric<br>• Thyme<br>• Oregano<br>• Rosemary<br>• Cloves<br>• Garlic<br>• Ginger<br>• Onions | Dried loose herbs<br>Salt-free herb mixes | Herb mixes containing salt<br>Herb preparations (commercial)<br>Essential oils derived from herbs |
|  | • Pepper<br>• Saffron<br>• Fennel<br>• Parsley |  |  |
| **Bread toppings (summary of the above)** | 20+ and 30+ cheeses (plain, with cumin, or with herbs)<br>Soft goat's cheese<br>Mozzarella<br>Dairy spread without additives<br>Cottage cheese (huttenkäse)<br>Boiled egg<br>Poached egg<br>Apple syrup (appelstroop) or 100% jam without additives or added sugar — preferably home-made<br>Home-made vegetable spread<br>Smoked salmon or smoked salmon trout<br>Salted herring, mackerel, eel<br>Honey<br>Vegetables<br>Fruit<br>100% peanut butter<br>100% nut spread | — | Chocolate sprinkles (hagelslag)<br>Chocolate flakes<br>Chocolate spread<br>Nut spread or peanut butter with added salt or sugar<br>Cheese substitutes<br>Processed deli meats<br>Sweet coconut bread topping (kokosbrood)<br>Fruit sprinkles (vruchtenhagel)<br>Aniseed-coated sugar sprinkles (muisjes) |

## Supplementary S4. Example weekly and daily menu

Two 6-week schedules were prepared for the dietary intervention. Participants were free to consume the meals on different days than scheduled, taking the best-before dates of the delivered food products into account. For the warm meals, the menu was designed to provide considerable variation. An example weekly menu for the warm meals is shown below.

### EXAMPLE WEEKLY MENU

| Day | Theme | Meal |
| --- | --- | --- |
| Monday | “Start the week without meat” | Vegetarian dish |
| Tuesday | “Local products” | Meal based on local products |
| Wednesday | “World meals” | International cuisine |
| Thursday | “Dutch delicious” | Potatoes / meat / vegetables |
| Friday | “Fish on Friday” | Meal with fish |
| Saturday | “Endless weekend” | Easy vegetarian meal |
| Sunday | “Lazy Sunday” | Easy and quick meal |

Each week includes:

- Fish twice a week
- A vegetarian meal twice a week
- Daily: at least 150 g of vegetables with the warm meal and 150 g of starch. The total daily amount of vegetables is 250 g, which may also be consumed at lunch or with snacks.
- Total composition of the warm meal: 500–700 kcal and >10 g fibre per person
- Olive oil used for preparation

### EXAMPLE DAILY MENU

#### Breakfast

- 2 slices of wholegrain bread with low-fat margarine (Dutch: halvarine) with added omega-3 fatty acids
- 1 slice with 20+ cheese and 1 slice with home-made apple syrup (Dutch: appelstroop) or 100% jam without added sugar
- 1 cup of buttermilk (Dutch: karnemelk) or kefir

#### Lunch

- 3 slices of wholegrain bread with low-fat margarine with added omega-3 fatty acids
- 1 slice with avocado or vegetable
- 1 slice with fruit
- 1 slice with 100% peanut butter or 100% nut spread
- 1 cup of buttermilk or kefir
- Water, (green) tea, or filter coffee without sugar or milk Dinner
- 125 g fish, poultry, or plant-based protein, prepared with olive oil
- 4 large spoonfuls of vegetables
- 3 potatoes, or spoonfuls of brown rice (Dutch: zilvervliesrijst) or wholegrain pasta
- 1 serving of plain low-fat or semi-skimmed yoghurt or quark (Dutch: kwark) with unsweetened dried fruit Snacks
- Water, (green) tea, or filter coffee without sugar or milk
- 2 servings of fruit (berries at least twice a week)

• 1 handful of unsalted nuts

• 1 serving of plain low-fat or semi-skimmed yoghurt or quark

